# Severe acute malnutrition promotes bacterial binding over pro-inflammatory cytokine secretion by circulating innate immune cells

**DOI:** 10.1101/2023.04.10.23288163

**Authors:** Tracy N. Phiri, Kuda Mutasa, Sandra Rukobo, Margaret Govha, Patience Mushayanembwa, Simutanyi Mwakamui, Tafhima Haider, Kanekwa Zyambo, Cherlynn Dumbura, Joice Tome, Thompson Runodamoto, Leah Chidamba, Florence D. Majo, Deophine Ngosa, Kanta Chandwe, Chanda Kapoma, Benjamin Mwapenya, Jonathan P. Sturgeon, Ruairi C. Robertson, Melanie Smuk, Robert Ntozini, Kusum Nathoo, Beatrice Amadi, Paul Kelly, Mutsa Bwakura-Dangarembizi, Andrew J. Prendergast, Claire D. Bourke

## Abstract

Children with severe acute malnutrition (SAM) are at high risk of infectious mortality and morbidity during and after hospital discharge. This risk persists despite nutritional and prophylactic antibiotic interventions among children with SAM, implicating persistent deficits in their immune defenses. Here we test the hypothesis that innate immune cells from children (0-59 months) hospitalized with SAM in Zambia and Zimbabwe (n=141) have distinct capacity to respond to bacteria relative to adequately-nourished healthy controls from the same communities (n=92). Neutrophils and monocytes from SAM inpatients had a higher capacity to bind *E. coli* but lower monocyte activation and pro-inflammatory mediator secretion in response to *E. coli* lipopolysaccharide (LPS) or heat-killed *Salmonella typhimurium* (HKST) than controls. Bacterial binding capacity differentiated children with SAM from controls after adjusting for clinical and demographic heterogeneity and normalized with duration of hospital treatment. Wasting severity, HIV status, and age group were associated with LPS and HKST-induced cytokine secretion, monocyte activation, and myeloperoxidase secretion, respectively. Bacterial binding capacity and monocyte activation during hospitalization were associated with higher odds of persistent SAM at discharge; a risk factor for subsequent mortality. Thus, SAM shifts anti-bacterial innate immune cell function, favoring bacterial containment over pro-inflammatory activation upon challenge, which contributes to persistent health deficits among hospitalized children.

**TEASER:** Children with severe acute malnutrition have distinct anti-bacterial innate immune cell function compared to healthy children which persists during their hospitalization and contributes to persistent wasting.

## MAIN TEXT

Wasting is a form of undernutrition that affects 6.7% of all children under 5 years old (45.4 million) globally, 89% of whom live in low or low-to-middle income countries (LMIC)(*1*). Severe acute malnutrition (SAM) is the most life-threatening form of wasting, clinically characterized as severe wasting (weight-for-height Z score (WHZ) ≤-3 and/or mid-upper arm circumference (MUAC) <115 mm, for children aged 6–59 months (mo)) with or without bilateral pitting edema(*1, 2*). Children with clinical complications of SAM requiring hospital admission (i.e. one of the Integrated Management of Childhood Illness clinical danger signs, acute infection, severe edema, or feeding problems(*1*)) are at particularly high risk of mortality; a meta-analysis of 19 studies among children under 5 years old conducted in 8 African countries found a mean all-cause mortality during hospitalization of 15.7% (95% confidence interval (CI): 10.4, 21.0)(*3*). Such high inpatient mortality rates and the persistent wasting that affects survivors of SAM, compromise ambitious global development goals to reduce global under 5 mortality, reduce and maintain childhood wasting to less than 5% by 2025 (3% by 2030), and achieve zero hunger(*4, 5*). Despite its definition according to anthropometry, it is increasingly recognized that complicated SAM is a combination of conditions rather than a single disorder(*3, 6–9*) with poor clinical outcomes before and after discharge from hospital(*6, 9–11*) and a pathophysiology that is not fully understood.

Pathogen carriage is common among children with complicated SAM at hospital admission(*8, 10, 12-16*) and empirical antibiotic treatment is therefore recommended even in the absence of infectious symptoms(*2*). Infections also worsen clinical outcomes among children with SAM: infections independently predict inpatient mortality(*3*), bacteremia is a frequent complication among children who die in hospital(*16*), and the risk of infectious mortality increases incrementally with the severity of wasting(*17*). However, pathogen carriage can be also be high among non-wasted children in LMIC(*18, 19*) and children with SAM appear susceptible to a range of pathogen species and types(*8, 10, 12-16*), suggesting that poor clinical outcomes among children with SAM reflect a reduced capacity to prevent and contain infection rather than infection carriage *per se*. Consistent with this hypothesis, children with both wasting and stunting have higher circulating levels of pathogen associated molecular patterns (PAMP), including bacterial endotoxin (also termed lipopolysaccharide, LPS) and flagellin(*18, 20–23*), which are thought to derive from both disseminated infections and from chronic translocation of commensal microbes, PAMP and microbial metabolites across their compromised gut barrier. Elevated systemic levels of pro-inflammatory biomarkers correspond with circulating PAMP levels in undernourished children(*24*), and this inflammatory state is associated with mortality among children with complicated SAM during hospitalization and after discharge(*15, 25, 26*), and positively correlated with metabolic disturbances associated with inpatient mortality(*26*). In other clinical conditions, such as gram-negative sepsis, systemic circulation of bacterial PAMP drives functional adaptation of circulating immune cells to limit lethal systemic inflammation(*27, 28*) but may also compromise defense against new infectious challenges(*29–32*). Multiple innate and adaptive immune mediators are dysregulated in undernourished children(*24, 33, 34*), indicative of an altered immune phenotype. However, few studies have directly assessed the capacity of immune cells to respond to new challenges (immune cell function) among cohorts of undernourished children adequately powered for their inherent clinical, demographic and immune heterogeneity; fewer still have evaluated how clinical characteristics of complicated SAM influence immune function relative to healthy controls from the same community(*24, 33, 34*).

In this study we evaluated how innate immune cells from children under 5 years old admitted to hospitals in Zambia and Zimbabwe with complicated SAM respond to bacterial PAMP challenge *in vitro*; a model for how each child might respond to a newly-acquired infection. We focused on gram-negative bacterial stimuli derived from species frequently identified in clinical isolates from children with complicated SAM (*Escherichia coli (E. coli)* and *Salmonella typhimurium (S. typhimurium)*)(*10, 15, 35*) and the two most abundant circulating innate immune cell types, monocytes and neutrophils. We addressed 3 outstanding questions in nutritional immunology: 1) how does the capacity to respond to bacterial challenge differ between innate immune cells from children hospitalized with SAM and adequately-nourished children? 2) Which clinically-relevant characteristics of children with SAM contribute to heterogeneity in their anti-bacterial innate immune cell function? 3) Does anti-bacterial innate immune cell function among children with SAM predict nutritional status at hospital discharge?

## RESULTS

### Demographic and clinical characteristics of the study cohort

We undertook anti-bacterial innate immune cell function assessment using blood samples from 141 children under 5 years old who were hospitalized with complicated SAM (defined according to WHO criteria(*1*)) and 92 adequately-nourished (WHZ>-1) children with no symptoms of infection (healthy controls; **Table 1**). All children were participants in a longitudinal observational cohort study of the health outcomes and pathogenesis of SAM (HOPE-SAM(*36*)) which recruited from 3 peri-urban tertiary hospital sites in Zambia (University Teaching Hospital, Lusaka) and Zimbabwe (Harare Central and Parirenyatwa Hospitals, Harare); inclusion of HOPE-SAM participants in this study is shown in **Figure S1.** Though all participants in the SAM group were critically unwell, they tended to be healthier (i.e. lower mortality, slightly older, less severe wasting and a lower percentage had persistent SAM at discharge) than those included in the wider HOPE-SAM cohort (**Table S1**).

**Table 1.**
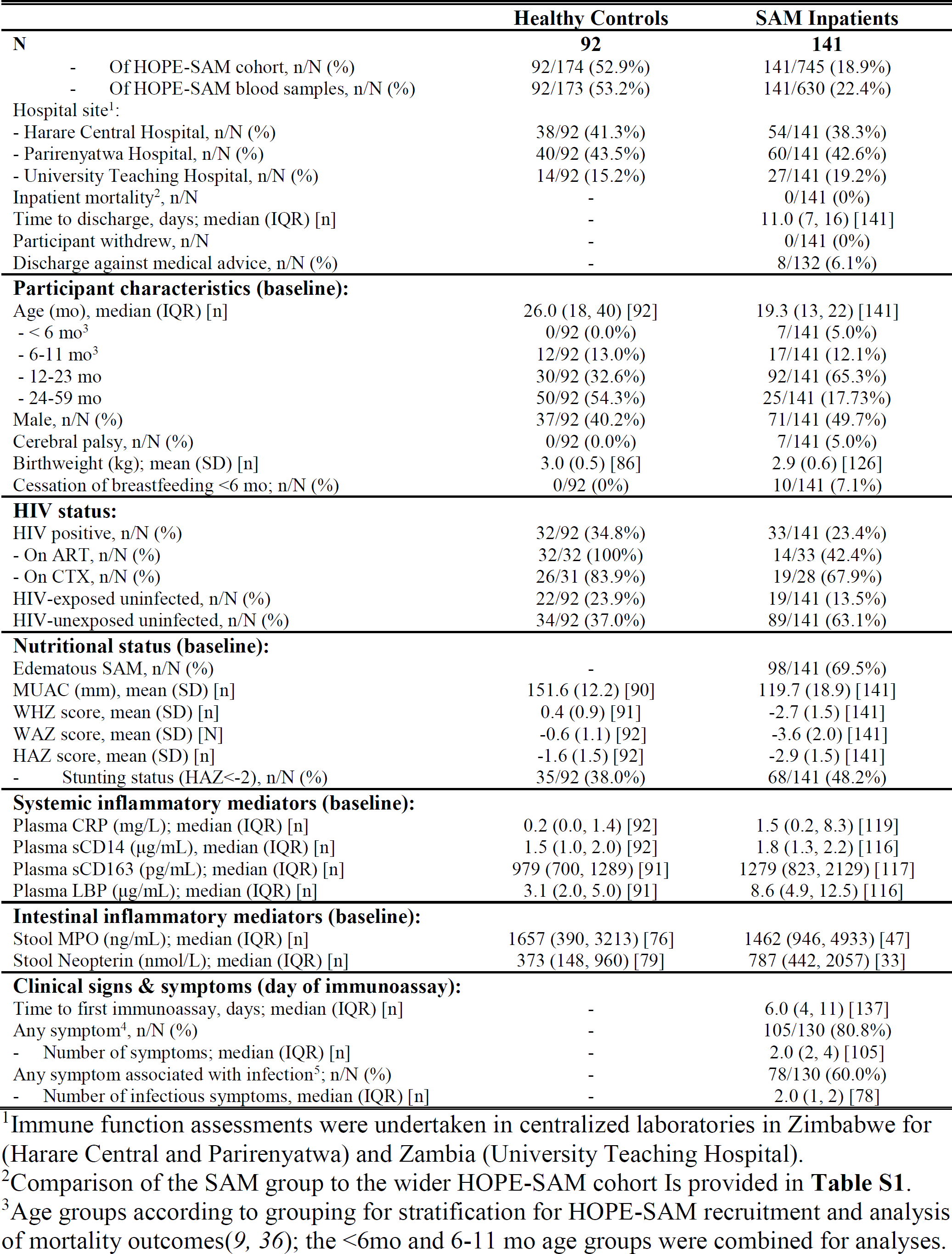
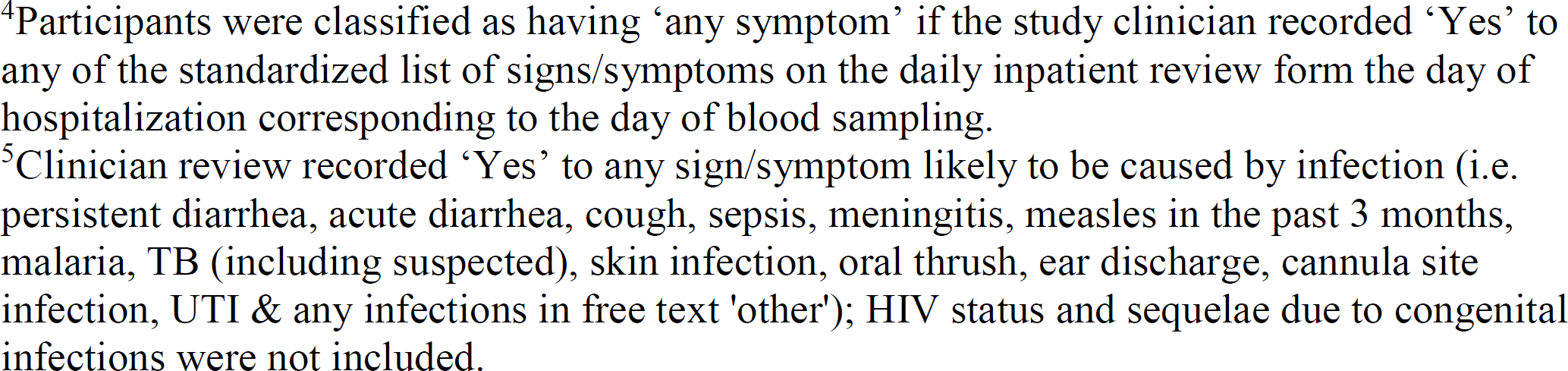
Summary of available data on baseline demographic and clinical characteristics of the SAM and Healthy control groups of the IMMUNO-SAM cohort

|  | Healthy Controls | SAM Inpatients |
| --- | --- | --- |
| **N** | **92** | **141** |
| - Of HOPE-SAM cohort, n/N (%) | 92/174 (52.9%) | 141/745 (18.9%) |
| - Of HOPE-SAM blood samples, n/N (%) | 92/173 (53.2%) | 141/630 (22.4%) |
| Hospital site<sup>1</sup>: |  |  |
| - Harare Central Hospital, n/N (%) | 38/92 (41.3%) | 54/141 (38.3%) |
| - Parirenyatwa Hospital, n/N (%) | 40/92 (43.5%) | 60/141 (42.6%) |
| - University Teaching Hospital, n/N (%) | 14/92 (15.2%) | 27/141 (19.2%) |
| Inpatient mortality<sup>2</sup>, n/N | - | 0/141 (0%) |
| Time to discharge, days; median (IQR) [n] | - | 11.0 (7, 16) [141] |
| Participant withdrew, n/N | - | 0/141 (0%) |
| Discharge against medical advice, n/N (%) | - | 8/132 (6.1%) |
| **Participant characteristics (baseline):** |  |  |
| Age (mo), median (IQR) [n] | 26.0 (18, 40) [92] | 19.3 (13, 22) [141] |
| - < 6 mo<sup>3</sup> | 0/92 (0.0%) | 7/141 (5.0%) |
| - 6-11 mo<sup>3</sup> | 12/92 (13.0%) | 17/141 (12.1%) |
| - 12-23 mo | 30/92 (32.6%) | 92/141 (65.3%) |
| - 24-59 mo | 50/92 (54.3%) | 25/141 (17.73%) |
| Male, n/N (%) | 37/92 (40.2%) | 71/141 (49.7%) |
| Cerebral palsy, n/N (%) | 0/92 (0.0%) | 7/141 (5.0%) |
| Birthweight (kg); mean (SD) [n] | 3.0 (0.5) [86] | 2.9 (0.6) [126] |
| Cessation of breastfeeding <6 mo; n/N (%) | 0/92 (0%) | 10/141 (7.1%) |
| **HIV status:** |  |  |
| HIV positive, n/N (%) | 32/92 (34.8%) | 33/141 (23.4%) |
| - On ART, n/N (%) | 32/32 (100%) | 14/33 (42.4%) |
| - On CTX, n/N (%) | 26/31 (83.9%) | 19/28 (67.9%) |
| HIV-exposed uninfected, n/N (%) | 22/92 (23.9%) | 19/141 (13.5%) |
| HIV-unexposed uninfected, n/N (%) | 34/92 (37.0%) | 89/141 (63.1%) |
| **Nutritional status (baseline):** |  |  |
| Edematous SAM, n/N (%) | - | 98/141 (69.5%) |
| MUAC (mm), mean (SD) [n] | 151.6 (12.2) [90] | 119.7 (18.9) [141] |
| WHZ score, mean (SD) [n] | 0.4 (0.9) [91] | -2.7 (1.5) [141] |
| WAZ score, mean (SD) [N] | -0.6 (1.1) [92] | -3.6 (2.0) [141] |
| HAZ score, mean (SD) [n] | -1.6 (1.5) [92] | -2.9 (1.5) [141] |
| - Stunting status (HAZ<-2), n/N (%) | 35/92 (38.0%) | 68/141 (48.2%) |
| **Systemic inflammatory mediators (baseline):** |  |  |
| Plasma CRP (mg/L); median (IQR) [n] | 0.2 (0.0, 1.4) [92] | 1.5 (0.2, 8.3) [119] |
| Plasma sCD14 (µg/mL), median (IQR) [n] | 1.5 (1.0, 2.0) [92] | 1.8 (1.3, 2.2) [116] |
| Plasma sCD163 (pg/mL); median (IQR) [n] | 979 (700, 1289) [91] | 1279 (823, 2129) [117] |
| Plasma LBP (µg/mL); median (IQR) [n] | 3.1 (2.0, 5.0) [91] | 8.6 (4.9, 12.5) [116] |
| **Intestinal inflammatory mediators (baseline):** |  |  |
| Stool MPO (ng/mL); median (IQR) [n] | 1657 (390, 3213) [76] | 1462 (946, 4933) [47] |
| Stool Neopterin (nmol/L); median (IQR) [n] | 373 (148, 960) [79] | 787 (442, 2057) [33] |
| **Clinical signs & symptoms (day of immunoassay):** |  |  |
| Time to first immunoassay, days; median (IQR) [n] | - | 6.0 (4, 11) [137] |
| Any symptom<sup>4</sup>, n/N (%) | - | 105/130 (80.8%) |
| - Number of symptoms; median (IQR) [n] | - | 2.0 (2, 4) [105] |
| Any symptom associated with infection<sup>5</sup>; n/N (%) | - | 78/130 (60.0%) |
| - Number of infectious symptoms, median (IQR) [n] | - | 2.0 (1, 2) [78] |

<sup>1</sup>Immune function assessments were undertaken in centralized laboratories in Zimbabwe for (Harare Central and Parirenyatwa) and Zambia (University Teaching Hospital).
<sup>2</sup>Comparison of the SAM group to the wider HOPE-SAM cohort Is provided in **Table S1**.
<sup>3</sup>Age groups according to grouping for stratification for HOPE-SAM recruitment and analysis of mortality outcomes(9, 36); the <6mo and 6-11 mo age groups were combined for analyses.
<sup>4</sup>Participants were classified as having ‘any symptom’ if the study clinician recorded ‘Yes’ to any of the standardized list of signs/symptoms on the daily inpatient review form the day of hospitalization corresponding to the day of blood sampling.
<sup>5</sup>Clinician review recorded ‘Yes’ to any sign/symptom likely to be caused by infection (i.e. persistent diarrhea, acute diarrhea, cough, sepsis, meningitis, measles in the past 3 months, malaria, TB (including suspected), skin infection, oral thrush, ear discharge, cannula site infection, UTI & any infections in free text 'other'); HIV status and sequelae due to congenital infections were not included.

Innate immune cell function was characterized in the SAM group (**Table 1**) using the first available venous blood sample (≥2mL) collected during their inpatient period (median time from admission to immune function assessment (time to immunoassay): 6.0 days, interquartile range (IQR): 4, 11); 38.2% of the SAM group had their first immune function assessment on the day of discharge from hospital. Clinical symptoms were recorded daily during hospitalization with 80.8% of children in the SAM group having ongoing clinical complications and 60.0% having symptoms indicative of acute infection at the time of immune function assessment (**Table S2**). All children who had immune function assessment during hospitalization survived to hospital discharge, spending a median of 11.0 (IQR: 7.0, 16.0) days in hospital. Healthy controls had a venous blood sample collected for immune function assessment at enrolment.

**Table 2.** Cross-fit partialling-out lasso regression of anti-bacterial innate immune function profiles (principal components) by demographic and al variables associated with inpatient and post-discharge mortality among children with SAM

|  |  | PC1<sup>1, 2</sup> | PC2<sup>1, 3</sup> | PC3<sup>1, 4</sup> | PC4<sup>1, 5</sup> | PC5<sup>1, 6</sup> |
| --- | --- | --- | --- | --- | --- | --- |
| **Baseline age group:** |  |  |  |  |  |  |
| - 0-11mo |  | *Reference* |  |  |  |  |
|  | Adj. Coef. | 0.02 | -0.73 | 0.49 | 0.39 | 0.12 |
| - 12-23mo | 95%CI | -1.19, 1.23 | -1.76, 0.29 | -0.39, 1.35 | -0.42, 1.20 | -0.50, 0.75 |
|  | P | 0.976 | 0.162 | 0.268 | 0.348 | 0.703 |
|  | Adj. Coef | -0.10 | -0.89 | 0.56 | 1.16 | 0.12 |
| - 24-59mo | 95%CI | -1.49, 1.28 | -2.15, 0.37 | -0.43, 1.55 | 0.24, 2.08 | -0.64, 0.89 |
|  | P | 0.885 | 0.165 | 0.265 | 0.013 | 0.753 |
| **HIV status:** |  |  |  |  |  |  |
| - Negative |  | *Reference* |  |  |  |  |
|  | Adj. Coef. | -0.77 | 0.46 | -0.66 | -0.23 | 0.55 |
| - Positive | 95%CI | -1.80, 0.26 | -0.45, 1.37 | -1.28, -0.04 | -1.00, 0.55 | -0.11, 1.21 |
|  | P | 0.143 | 0.322 | 0.037 | 0.566 | 0.101 |
| **Baseline edema status:** |  |  |  |  |  |  |
| - Non-edematous |  | *Reference* |  |  |  |  |
|  | Adj. Coef. | 0.51 | 0.12 | 0.11 | 0.47 | -0.09 |
| - Edematous | 95%CI | -0.59, 1.61 | -0.87, 1.12 | -0.52, 0.75 | -0.17, 1.11 | -0.64, 0.45 |
|  | P | 0.363 | 0.809 | 0.723 | 0.152 | 0.741 |
| **Symptom/s of infection at time of immunoassay:** |  |  |  |  |  |  |
| - None |  | *Reference* |  |  |  |  |
|  | Adj. Coef. | 0.28 | 0.33 | -0.10 | -0.07 | 0.10 |
| - ≥1 symptom | 95%CI | -0.62, 1.17 | -0.43, 1.09 | -0.63, 0.43 | -0.59, 0.46 | -0.36, 0.55 |
|  | P | 0.545 | 0.395 | 0.712 | 0.799 | 0.684 |
|  | Adj. Coef. | -0.02 | 0.04 | 0.00 | 0.00 | 0.00 |
| **Baseline MUAC (mm)** | 95%CI | -0.06, 0.03 | 0.01, 0.07 | -0.02, 0.02 | -0.02, 0.02 | -0.03, 0.03 |
|  | P | 0.427 | 0.010 | 0.834 | 0.690 | 0.860 |

<sup>1</sup>n=109 children with SAM; fixed exposure variables selected based on HOPE-SAM mortality and morbidity outcomes(6, 9); controls offered to model: sex, hospital site, time to immunoassay, immune function assessment on the day of hospital discharge, baseline WHZ, baseline HAZ, birthweight & cerebral palsy status
<sup>2</sup>5 controls, Wald chi<sup>2</sup>: 3.20, p=0.783
<sup>3</sup>3 controls, Wald chi<sup>2</sup>: 9.45, p=0.150
<sup>4</sup>3 controls, Wald chi<sup>2</sup>: 7.77, p=0.255
<sup>5</sup>3 controls, Wald chi<sup>2</sup>: 13.74, p=0.033
<sup>6</sup>4 controls, Wald chi<sup>2</sup>: 3.33, p=0.760

33 children in the SAM group (23.4%) and 32 children in the healthy control group (34.8%) were living with HIV. 42.4% of HIV-positive children in the SAM group were on antiretroviral therapy (ART) at hospital admission and 100% of HIV-positive children in the healthy control group were on ART.

Consistent with previous studies(*24, 33, 34*), children with complicated SAM had higher concentrations of systemic and intestinal inflammatory biomarkers relative to healthy controls (**Table 1**); these included elevated monocyte/macrophage-derived surface marker levels (sCD14, sCD163), acute phase proteins (CRP) and LPS-binding protein (LBP) in plasma and innate immune cell granules in stool (MPO and neopterin).

### SAM enhances the capacity of innate immune cells to bind bacteria

To quantify the capacity of blood immune cells to bind to bacteria, we incubated whole blood samples from each child either without stimulus (negative control) or with *E. coli*-coated bioparticles labelled with a fluorescent dye (AlexaFluor 488, AF488) at 37⁰C for 1h and analyzed cell-type specific binding to *E. coli* via flow cytometry (**Figure S2**). This assay identifies cells that have either bound or internalized the bioparticles and therefore reflects both bacterial binding and phagocytic capacity. The SAM and healthy control groups had similar total leukocyte counts, and percentages of lymphocytes (positive for any lymphocyte lineage marker (Lin): CD3, CD19, CD20 and/or CD56) and neutrophils (Lin-CD66b+CD16+) in unstimulated cultures (**Table S3**; **Figure S3**). Percentages of monocytes (Lin-CD66b-HLA-DR+) were slightly lower in unstimulated cultures from the SAM group (median: 2.3%; IQR: 1.3, 4.0) versus healthy control group (3.7%, IQR: 2.7, 5.3; p=0.004) with lower percentages of classical monocytes (CD14^hi^CD16-; median: 30.6% (IQR: 16.2, 41.0) versus 35.5% (IQR: 26.4, 46.4); p=0.025), higher percentages of intermediate monocytes (CD14^hi^CD16+; median: 12.0% (IQR: 7.8, 16.0) versus 8.3% (IQR: 5.9, 11.9); p<0.001) and similar percentages of non-classical monocytes (CD14^lo^CD16+; median: 29.8% (IQR: 18.1, 40.1) versus 32.7% (IQR: 22.7, 41.2); p=0.410). The classical, intermediate and non-classical monocyte sub-sets reflect a maturation continuum(*37*), expanded intermediate monocytes therefore reflect increased circulating monocyte activation and maturation in SAM versus healthy controls. Unstimulated cultures for both groups had negligible *E. coli*+ events and AF488 meanFIs (**Figure S2B; Table S4**).

After incubation with *E. coli*-bioparticles, there were higher median percentages of *E. coli*+ leukocytes in the SAM versus healthy control group (**Figure 1A**); evaluation of standardized numbers of *E. coli*+ leukocytes from each participant (500 events/participant) via t-distributed stochastic neighbor embedding (tSNE; **Figure 1B**) and clustering of *E.coli*+ cell types according to their cell surface marker expression via flow cytometry self-organizing maps (FlowSOM; **Figure S4**) identified compositional differences in the immune cell types that bound bacteria in samples from the SAM versus healthy control group. There was no difference in total leukocyte count (**Figure 1C**), however *E. coli*+ leukocyte count (**Figure 1D**) and percentage of *E. coli*+ leukocytes (**Figure 1E**) were higher in the SAM versus healthy control group; analyses are summarized in **Table S4.** *E. coli* meanFI of total leukocytes were lower in the SAM group relative to healthy controls (**Figure 1F**), indicative of lower mean amounts of bioparticles bound per-cell. Neutrophils made up a higher percentage, lymphocytes a lower percentage and monocytes a similar percentage of *E. coli*+ leukocytes in the SAM versus healthy control group (**Figure 1G**). Within the monocyte population, classical monocytes made up a lower percentage of *E. coli*+ monocytes and the more differentiated intermediate monocyte phenotype made up a higher percentage of *E. coli*+ monocytes in the SAM versus healthy control group (**Figure 1H**).

**Figure 1.**
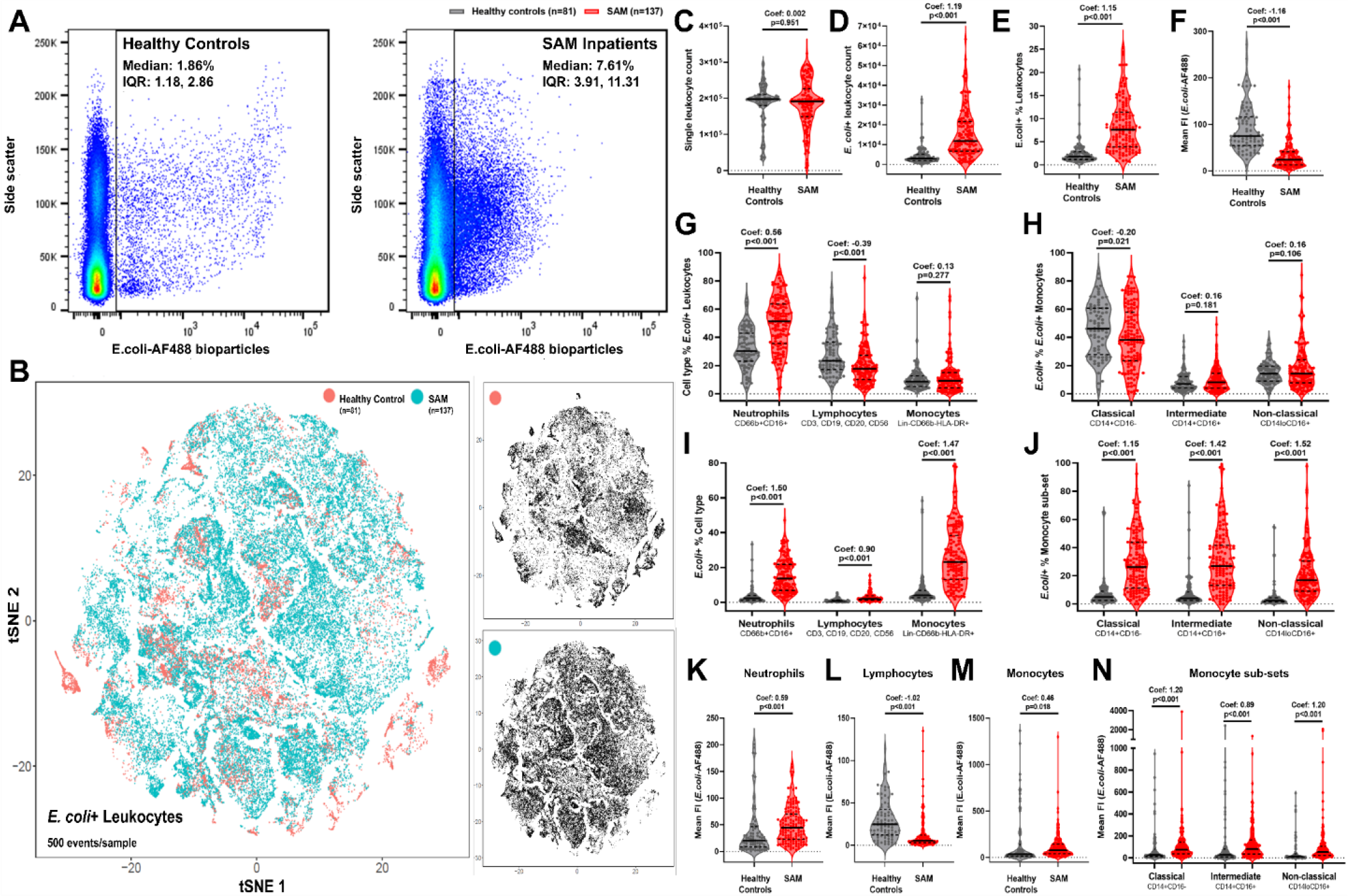
Circulating neutrophils and monocytes from children with SAM have enhanced bacterial binding capacity compared to healthy controls. A) Representative flow cytometry of bacterial binding by all single leukocytes from a healthy control (left) relative to a SAM inpatient (right) with median and IQR for each group. B) tSNE plot of 500 *E. coli*+ single leukocytes per sample clustered by cell surface markers (Lin, CD66b, CD14, CD16 and HLA-DR) colour-coded by healthy control (pink) and SAM (blue) group; plots separated by group (right). C) Total single leukocyte count, D) total *E. coli*+ single leukocyte count, E) total *E. coli*+ leukocytes as a percentage of all single leukocytes, and F) mean AF488 FI of all single leukocytes from flow cytometry analysis of healthy controls (grey, n=81) versus SAM inpatients (red; n=137). G) Leukocyte sub-types as a percentage of total *E. coli*+ leukocytes, and H) monocyte sub-sets as a percentage of total *E. coli*+ monocytes. I) *E. coli*+ cells as a percentage of all cells of that type, and J) *E. coli*+ monocyte sub-sets as a percentage of all cells of the sub-set. Mean AF488 FI of K) neutrophils, L) lymphocytes, M) monocytes and N) monocyte sub-sets. Violin plots indicate median (dark line) and IQR (dashed lines); coefficients (Coef.) and p-values are reported for unadjusted univariable negative binomial regression of event counts (C, D), linear regression of mean AF488 FI (F, K-N), and fractional regression of proportions (E, G-J) by SAM status.

To evaluate the cell-type specific capacity for *E. coli* binding, independent of compositional differences in the cell types present in the initial unstimulated blood sample, we compared *E. coli*+ percentages of lymphocytes, neutrophils, monocytes and monocyte sub-sets between groups. A higher percentage of all 3 cell types (**Figure 1I**) and all monocytes sub-sets (**Figure 1J**) bound to *E. coli* in the SAM group compared to the healthy control group, indicative of a generalized enhancement of cell-type specific bacterial binding capacity in the context of SAM which was more pronounced in neutrophils (fractional regression coefficient: 1.50, 95% CI: 1.19, 1.81; p<0.001) and monocytes (coefficient: 1.47, 95% CI: 1.15, 1.79; p<0.001) than in lymphocytes (coefficient: 0.90, 95% CI: 0.65, 1.15; p<0.001). Neutrophils from the SAM group had higher AF488 meanFI (**Figure 1K**), lymphocytes had lower meanFI (**Figure 1L**), and total monocytes (**Figure 1M**) and all monocyte sub-sets had higher meanFI (**Figure 1N**) than those from the healthy control group. Thus, the lower meanFI for total leukocytes in the SAM group (**Figure 1F**; >50% of leukocytes in both groups; **Figure S3**) reflected lower bacterial binding per-lymphocyte in the SAM group despite their higher bacterial binding by neutrophils and monocytes.

### SAM reduces monocyte activation and pro-inflammatory cytokine secretion in response to bacteria

Upregulation of major histocompatibility complexes (e.g. HLA-DR) and co-receptors (e.g. CD86 and CD40) required for activation of T cells, and secretion of soluble mediators to mobilize anti-bacterial functions in other cells and tissues, recruit immune cells, and promote bactericidal reactive oxygen species (ROS) production are key features of the innate immune response to bacterial infection. We quantified these characteristics in immune cells and supernatants from parallel 24h whole blood cultures conducted without stimulus (negative controls) and with bacterial PAMP (*E. coli* LPS and heat-killed *S. typhimurium* (HKST)).

Unstimulated whole blood cultures from children with SAM had higher percentages of neutrophils, and a more mature monocyte phenotype than those from healthy controls (**Figure S5A-B; Table S3**). Unstimulated monocytes from children with SAM expressed lower levels of HLA-DR (medianFI) than healthy controls but similar levels of CD86 and CD40 (**Figure S5C-E; Table S5**). Basal concentrations of pro-inflammatory cytokines (IL-6, IL-8 and TNFα) in unstimulated whole blood culture supernatants were similar in the two groups (**Figure S5F; Table S5**). Basal concentrations of myeloperoxidase (MPO), an enzyme which catalyzes production of ROS, were higher in culture supernatants from the SAM versus healthy control group (**Figure S5F; Table S5**).

To account for unstimulated differences between the two groups and focus on the capacity of immune cells to respond to a new bacterial challenge *in vitro*, we calculated PAMP-induced cell surface marker expression and mediator secretion for each child by subtracting HLA-DR, CD86 and CD40 medianFIs and mediator concentrations present in their corresponding unstimulated cultures (Δ); Δmedian FIs/concentrations were censored at zero (i.e. no increase in response to PAMP). Monocytes from children with SAM had lower LPS-induced ΔmedianFI for HLA-DR, CD86 and CD40 than those from healthy controls (**Figure 2A-D**; **Table S5**). The same pattern was observed for HKST-induced monocyte surface marker ΔmedianFI but with stronger evidence for a difference in HKST-induced CD40 than for HKST-induced HLA-DR and CD86 (**Figure 2B-D**; **Table S5**). LPS- and HKST-induced IL-6, IL-8, TNFα and MPO were all lower in supernatants from cultures conducted with blood samples from SAM versus healthy controls (**Figure 2E-F**; **Table S5**).

**Figure 2.**
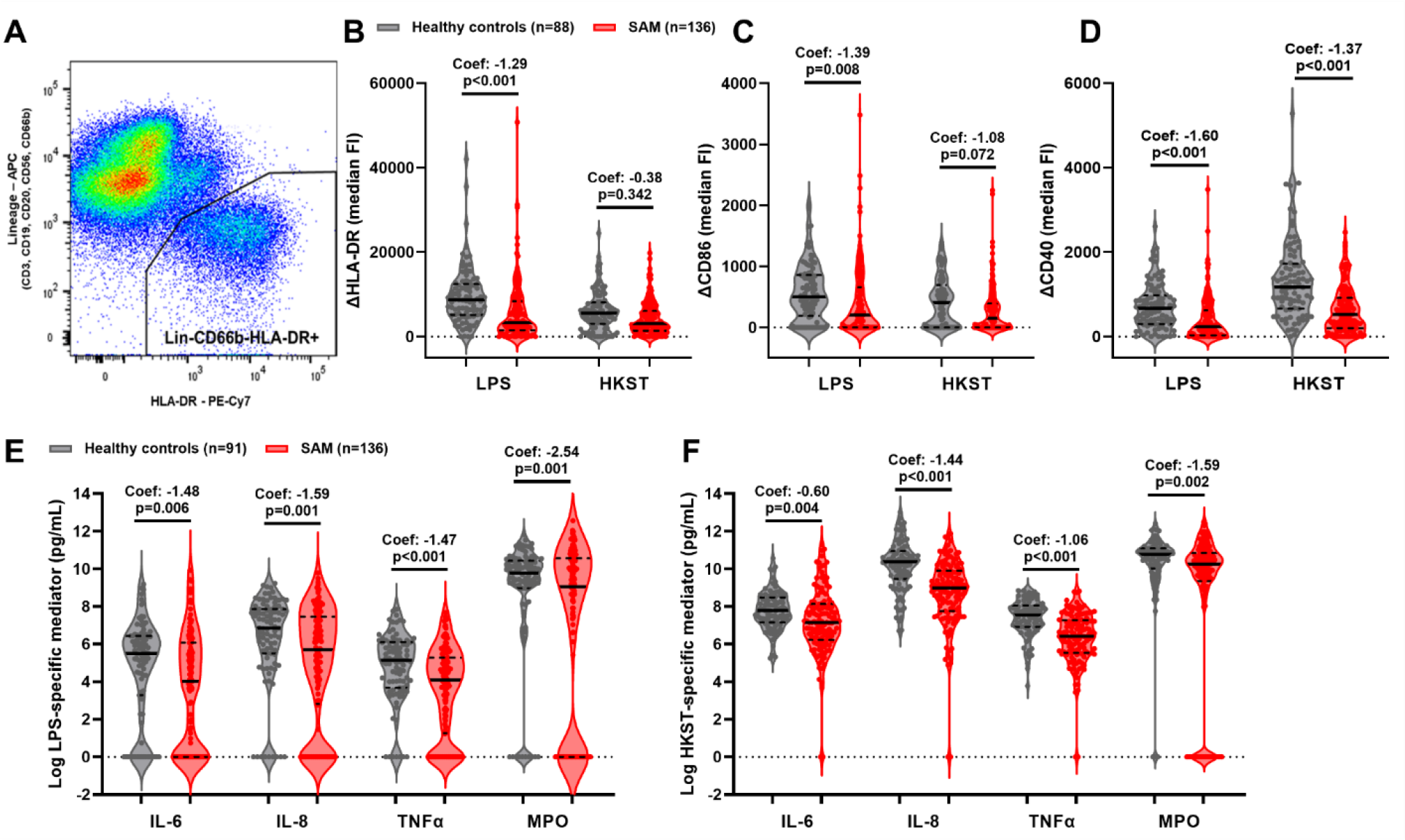
SAM compromises monocyte maturation and pro-inflammatory mediator secretion in response to bacterial PAMP. Whole blood samples from children with SAM (red) and healthy controls (grey) were incubated for 24h in media alone, LPS or HKST; cultured cells were analysed by flow cytometry (A-D; Healthy controls, n=88; SAM, n=136) and culture supernatants were analysed by ELISA (E-F; Healthy controls, n=91; SAM, n=136). A) Flow cytometry gating of total monocytes after exclusion of cells expressing markers of lymphocyte and granulocyte lineage; representative example. PAMP-induced median FI of B) ΔHLA-DR-PECy7, C) ΔCD86-FITC, and D) ΔCD40-PerCP-Cy5.5 of total monocytes after subtraction of median FI in unstimulated cultures. E) LPS-induced and F) HKST-induced ΔIL-6, ΔIL-8, ΔTNFα and ΔMPO after subtraction of concentrations in unstimulated cultures. Violin plots indicate median (dark line) and IQR (dashed lines); coefficients (Coef.) and p-values are reported for unadjusted tobit regression by group.

### Monocytes and neutrophils from children with SAM prioritize bacterial containment over systemic immune activation

Having identified differences in individual read-outs of anti-bacterial innate immune cell function between the SAM and healthy control groups, we sought to characterize patterns of neutrophil and monocyte function that explained the immune heterogeneity in our cohort via principal components analysis (PCA) of all read-outs. 126/141 children (89.4%) in the SAM group and 73/92 children (79.3%) in the healthy control group had complete data for all variables which were included in a single PCA. We identified 5 groups of anti-bacterial monocyte and neutrophil functions (principal components, PC; **Figure 3A**): PC1 scores for each participant corresponded to their neutrophil and monocyte bacterial binding capacity; PC2 scores to theirs LPS- and HKST-induced pro-inflammatory cytokine (TNFα, IL-6 and IL-8) release; PC3 scores to their LPS- and HKST-induced monocyte maturation marker expression; PC4 scores to their LPS- and HKST-induced MPO; and, PC5 scores to a combination of their neutrophil bacterial binding and PAMP-induced MPO release (**Figure 3A**). PC1-5 collectively captured 72.1% of the variance in our immune function data.

**Figure 3.**
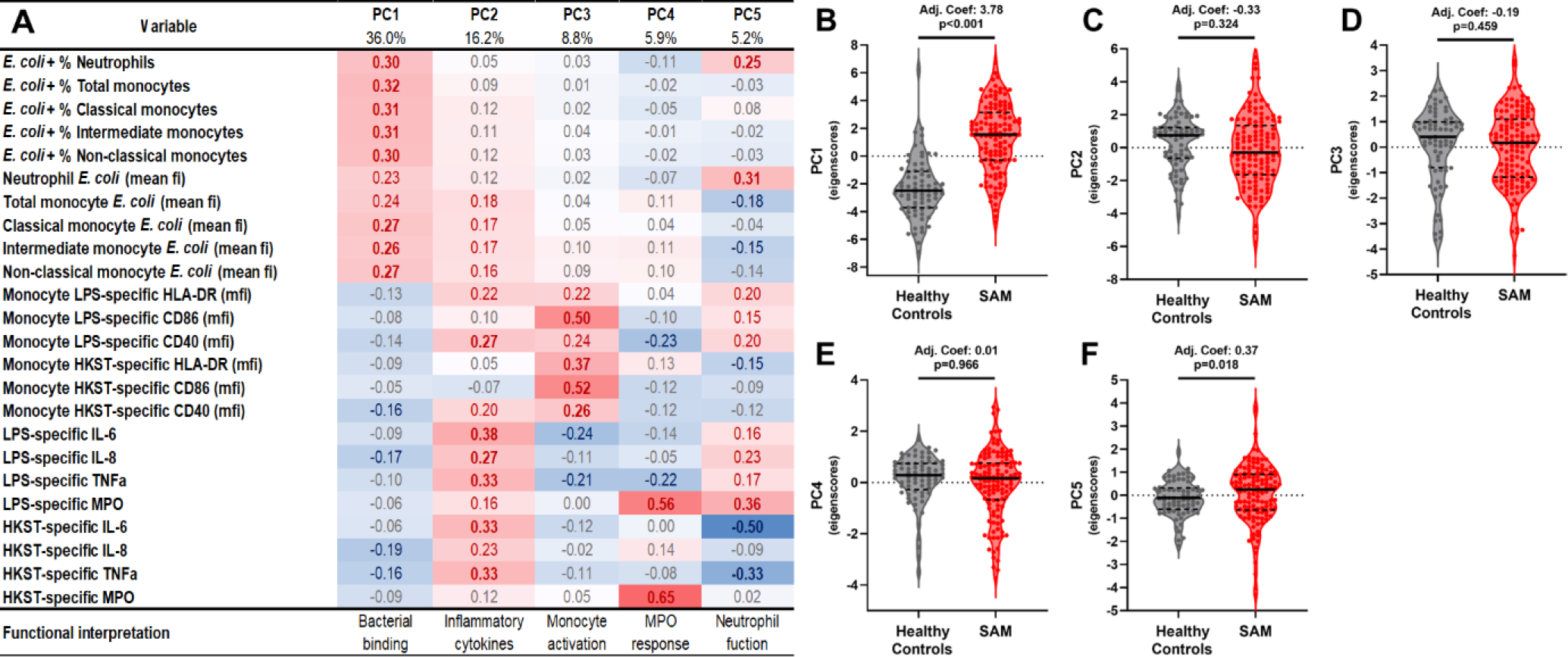
Monocytes and neutrophils of children with SAM prioritise bacterial containment over systemic immune activation. A) Factor loadings for all anti-anti-bacterial innate immune function variables included in a principal components analysis (126 children with SAM and 73 healthy controls with full data). Table colour-coded by strength and direction of association between the original variables and the resulting principal components (PC) with red indicating positive (bold for coefficient ≥0.3) and blue negative (bold for coefficient ≤-3) association. Percentages correspond to the amount of variance in the dataset accounted for by each PC. Functional interpretation reflects the combination of the original variables captured by each PC. Scores for B) PC1, C) PC2, D) PC3, E) PC4 and F) PC5 for each child in the SAM (red) and healthy control (grey) groups. Violin plots indicate median (dark line) and IQR (dashed lines); coefficients (Coef.) and p-values are reported for multivariable linear regression by SAM status, adjusting for sex, age group, HIV status, hospital site and immunoassay on the day of discharge.

We then tested our hypothesis that anti-bacterial innate immune cell function differs between children admitted to hospital with SAM relative to healthy controls from the same community using linear regression of participant scores for each PC, adjusted for characteristics that might affect both their nutritional status and their immune function (i.e. age group, sex, HIV status, hospital site, and assessment of immune function on the same day as hospital discharge; **Figure 3B-F; Table S6**). These analyses provide evidence for a total effect of SAM status on innate immune cell bacterial binding capacity (PC1 scores; adjusted coefficient: 3.69, 95%CI: 2.97, 4.40, p<0.001) and neutrophil anti-bacterial response (PC5 scores; adjusted coefficient: 0.33, 95%CI: 0.03, 0.63, p=0.031), consistent with a shift in innate immune cell function favoring containment of bacteria by binding/phagocytosis. Despite having higher circulating and intestinal pro-inflammatory biomarker levels relative to healthy controls (**Table 1**) and evidence that critically unwell children hospitalized in LMIC may have enhanced LPS-induced pro-inflammatory cytokine secretion(*38*), there was limited evidence that PC2 scores (adjusted coefficient: -0.48, 95%CI: -1.13, 0.16, p=0.143), PC3 scores (adjusted coefficient: -0.23, 95%CI: -0.73, 0.27, p=0.358) or PC4 scores (adjusted coefficient: 0.01, 95%CI: -0.39, 0.38, p=0.031) differed by nutritional status after adjusting for confounders (**Figure 3C-E**).

Since HIV status can influence many aspects of SAM and has a profound impact on the immune system, we undertook sensitivity analysis including only the HIV-negative children (**Figure 4A-E**; **Table S7**; SAM n=101/126 (80.2%), healthy controls n=46/73 (63.0%)); this strengthened the association between SAM status and PC1 (adjusted coefficient: 4.04, 95%CI: 3.17, 4.91, p<0.001) and PC5 (adjusted coefficient: 0.41, 95%CI: 0.07, 0.76, p=0.017). Furthermore, because some of our SAM group had immune function assessments late in the course of their inpatient rehabilitation, we undertook sensitivity analyses only including children who had their immune function assessed prior to the day of discharge (**Figure 4F-J; Table S8**; SAM n=75/126 (59.5%), healthy controls n=73/73 (100%)); this slightly weakened the effect size but did not affect the direction or significance of the association between SAM and PC1 scores (adjusted coefficient: 3.80, 95%CI: 3.09, 4.52, p<0.001) and PC5 scores (adjusted coefficient: 0.36, 95%CI: 0.05, 0.66, p=0.021). There remained limited evidence to support an association between SAM status and PC2-4 scores after adjusting for confounders in either sensitivity analysis (**Figure 4B-D** and **G-I**).

**Figure 4.**
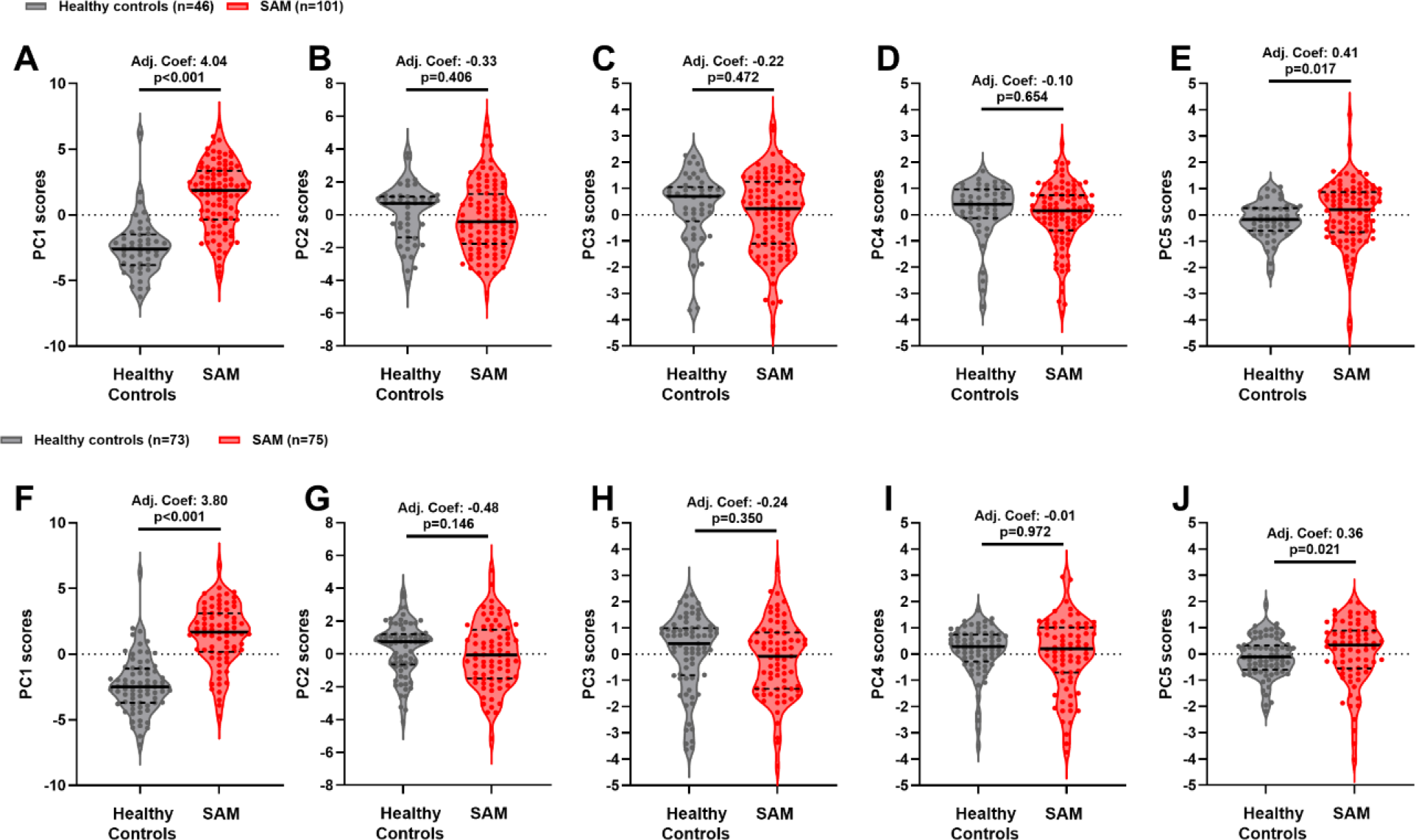
SAM differentiates innate anti-bacterial immune cell function from healthy controls among HIV-negative children and among children who had immune assessment prior to the day of discharge. Scores for A) PC1, B) PC2, C) PC3, D) PC4 and E) PC5 for HIV negative children in the healthy control (grey) and SAM (red) groups; coefficients (Coef.) and p-values are reported for multivariable linear regression by SAM status, adjusting for variation due to sex, age group, hospital site and immunoassay on the day of discharge. Scores for F) PC1, G) PC2, H) PC3, I) PC4 and J) PC5 for HIV for healthy controls (grey) and children in the SAM group for whom an immunoassay was conducted prior to the day of hospital discharge; coef. and p-values are reported for multivariable linear regression by SAM status, adjusting for variation due to sex, age group, HIV status and hospital site.

### Anti-bacterial innate immune cell function is partially shaped by age group, HIV status and wasting severity at hospital admission

Children with complicated SAM experience a range of comorbidities and risk factors for adverse clinical outcomes(*3, 6–9*), which could plausibly impact their immune function relative to healthy controls. To explore this hypothesis, we used cross-fit partialling-out lasso linear regression to estimate the effect of variables associated with mortality in the HOPE-SAM cohort (i.e. HIV status, nutritional edema, age group, MUAC at baseline, and symptoms of infection at the time of immune function assessment(*6, 9*)) on PC1-5 scores of the SAM group. Other potential sources of immune heterogeneity in the SAM group were offered to the model as control variables (i.e. sex, hospital site, time to immunoassay (days), immune assessment on the day of discharge, height-for-age Z score (HAZ), WHZ, cerebral palsy status, and birthweight (g)). 109/126 (86.5%) of children had complete data on all immune function clinical and demographic variables for these analyses.

We did not find evidence to support an independent effect of clinically-important variables on PC1 or PC5 scores (**Table 2**). However, we found evidence to support a positive association between PC2 scores and baseline MUAC (adjusted coefficient: 0.04, 95%CI: 0.01, 0.07, p=0.010; **Figure 5A**), a negative association between PC3 scores and HIV infection (adjusted coefficient: -0.66, 95%CI: -1.28, -0.04, p=0.037; **Figure 5B**), and a positive association between PC4 scores and age group; children in the 12-23mo (adjusted coefficient: 0.39, 95%CI: -0.42, 1.20, p=0.348) and 24-59mo (adjusted coefficient: 1.16, 95%CI: 0.24, 2.08, p=0.013) age groups had higher PC4 scores than the youngest age group (0-11mo; **Figure 5C**). The best evidence that clinically important exposure variables influenced anti-bacterial innate immune cell function came from the PC4 model (Wald chi^2^: 13.74, p=0.033) with weaker evidence from models for the other PCs (**Table 2**).

**Figure 5.**
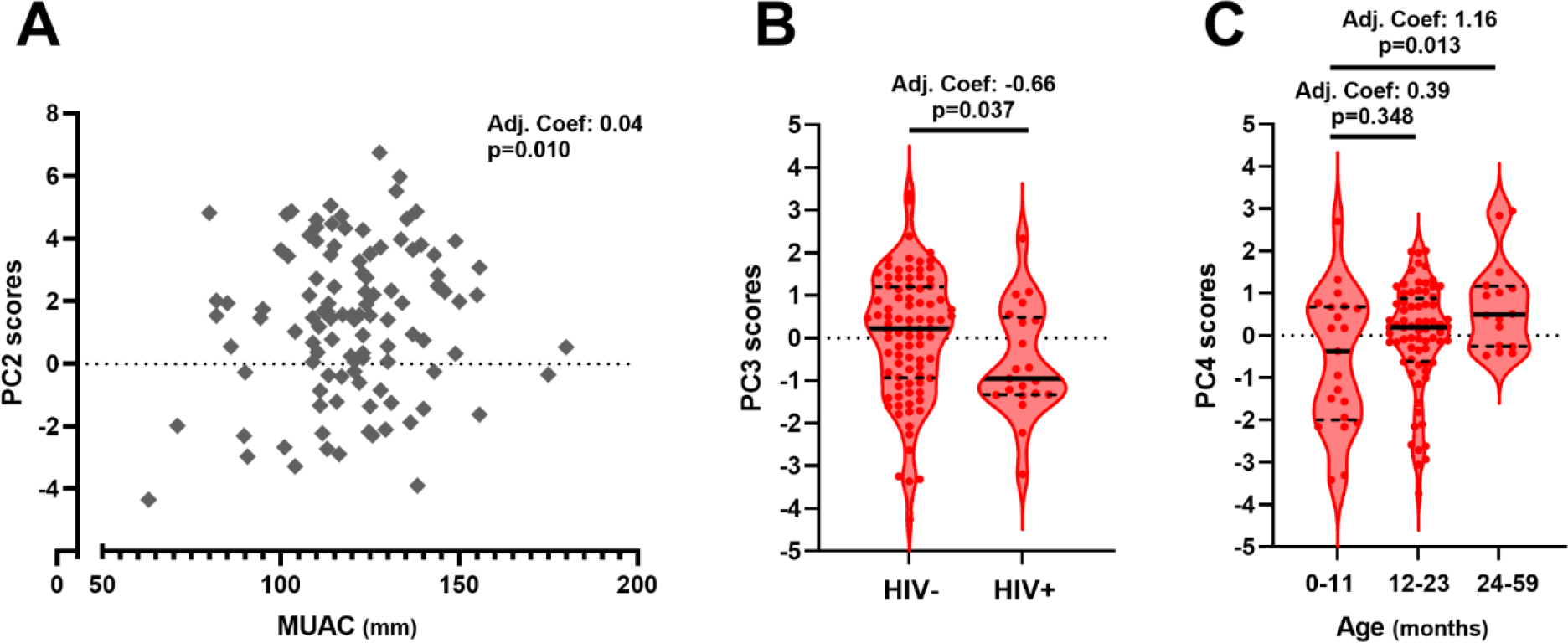
Clinical and demographic variables associated with adverse clinical outcomes from SAM shape some anti-bacterial innate immune cell functions. A) PC2 (anti-bacterial inflammatory cytokines) scores plotted against MUAC (mm), B) PC3 (anti-bacterial monocyte activation) scores plotted by HIV status, and C) PC4 (anti-bacterial MPO) plotted by participant age group for the SAM group (n=109). Coefficients (Coef.) and p-values are reported for multivariable cross-partialling out lasso linear regression of PC scores by clinically-relevant patient characteristics, adjusting for other clinically-relevant variables (baseline oedema, and presence of any symptom of infection at the time of immune function assessment) and controls. Only exposure variables where there was evidence for an effect on immune function are shown; full analysis is provided in Table 2.

### Anti-bacterial innate immune cell function normalizes with duration of inpatient rehabilitation

Inpatient rehabilitation of children with complicated SAM aims to resolve clinical complications, including acute infections, and begin nutritional stabilization and recovery so that children can be discharged to outpatient community management of SAM(*1*). Given that all children in the SAM group survived to discharge, we hypothesized that those who had a longer period of inpatient rehabilitation prior to their immune function assessment (time to immunoassay) would have innate immune cell function more similar to healthy controls than those assessed at hospital admission. We found cross-sectional evidence for a negative association between PC1 (**Figure 6A**; adjusted coefficient: -0.10, 95%CI: -0.15, -0.05, p<0.001) and PC5 (**Figure 6E**; adjusted coefficient: -0.03, 95%CI: -0.05, 0.01, p=0.040) and time to immunoassay after adjusting for hospital site, symptoms of infection on the day of immunoassay, and immune function assessment on the day of discharge (**Table S9**). We did not find substantial evidence that anti-bacterial pro-inflammatory cytokine secretion (PC2; adjusted coefficient: -0.01, 95%CI: -0.06, 0.04, p=0.784), upregulation of monocyte activation markers in response to bacterial PAMP (PC3; adjusted coefficient: 0.00, 95%CI: - 0.04, 0.04, p=0.981) or PAMP-induced MPO production (PC4; adjusted coefficient: 0.02, 95%CI: -0.02, 0.06, p=0.295) changed with time to immunoassay (**Figure 6B-D**; **Table S9**).

**Figure 6.**
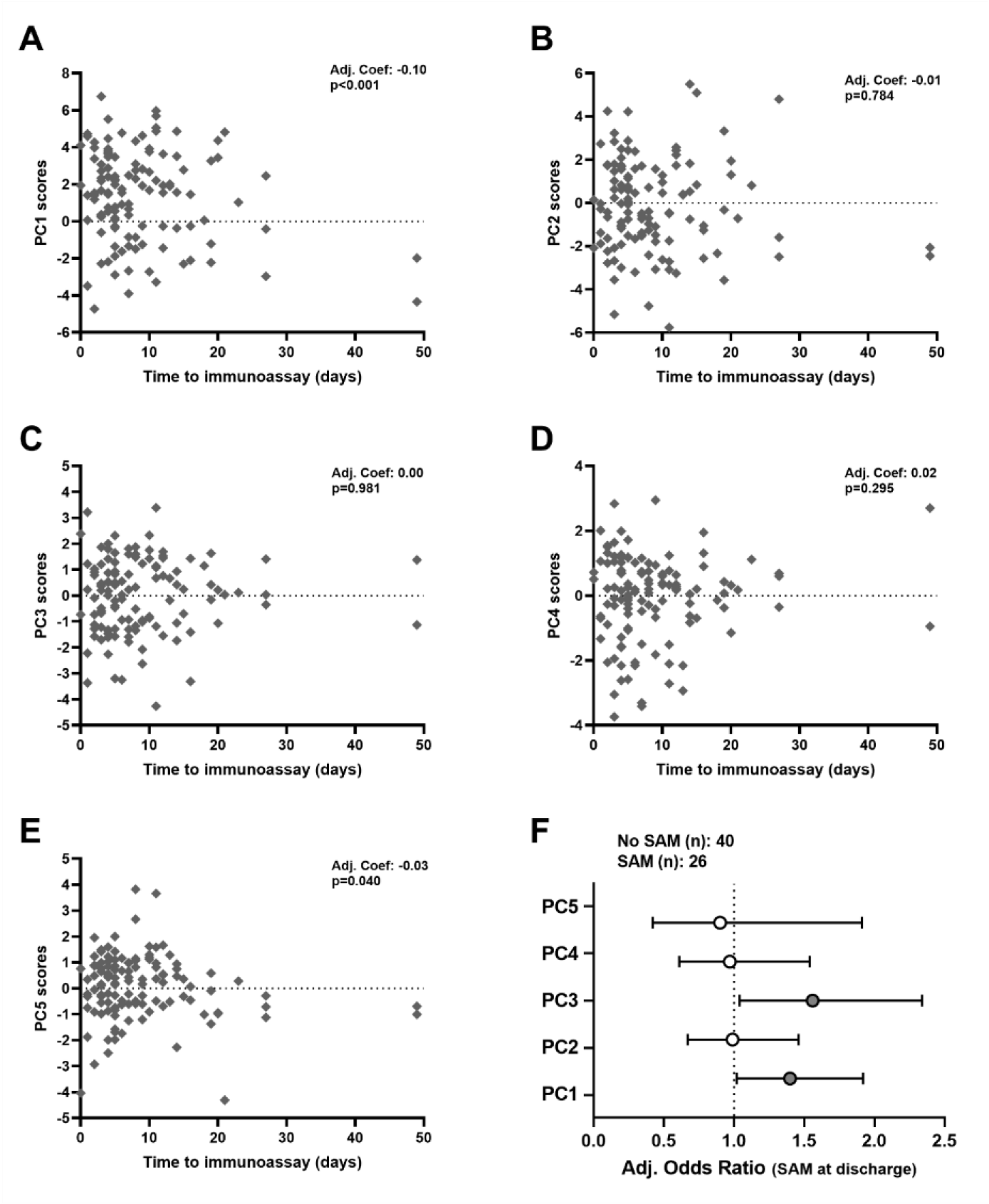
Anti-bacterial innate immune cell function of children with SAM partially normalises during inpatient rehabilitation and is associated with persistent SAM at hospital discharge. Scores for A) PC1, B) PC2, C) PC3, D) PC4 and E) PC5 for children with SAM (n=120) plotted against the time between their initial clinical assessment at hospital admission and immune function assessment (days); coefficients (Coef.) and p-values are reported for multivariable linear regression by time to immunoassay, adjusted for hospital site, symptom/s of infection on day of blood sampling, and immunoassay on the day of discharge. F) Odds ratios for persistent SAM at discharge among children who had immune function assessment prior to the day of discharge (n=66); logistic regression, adjusted for variation due to sex, age group at baseline, HIV status, hospital site and baseline MUAC.

### Anti-bacterial immune function during hospitalization is associated with persistent SAM at discharge

In the HOPE-SAM cohort, children with persistent SAM at discharge were at greater risk of subsequent mortality and had persistently lower WHZ and MUAC throughout 1 year of post-discharge follow-up than children who were discharged with moderate acute malnutrition (MAM) or adequate nutrition(*9*). In light of our observations that children with SAM had a distinct bacterial binding capacity to healthy controls, and cross-sectional evidence that these differences may normalize during inpatient rehabilitation, we hypothesized that immune function would be associated with persistent SAM at hospital discharge. We tested this hypothesis in children with SAM who had immune function assessment prior to the day of discharge and who were not discharged against medical advice (n=66); 26/66 (39.4%) had persistent SAM and 40/66 (60.6%) had MAM (-2<WHZ<-1; n=13) or adequate nutritional status (WHZ>-1; n=27) at discharge (**Table S10**).

Unadjusted logistic regression provided limited evidence for anti-bacterial innate immune function (PC1-5) affecting odds of persistent SAM at discharge (**Figure S6; Table S11**). However, after adjusting for confounders (sex, hospital site, HIV status, baseline age group and baseline MUAC), higher scores for both PC1 (bacterial binding capacity) and PC3 (monocyte activation) were associated with 1.40 (95% CI: 1.02, 1.92; p=0.037) and 1.56 (95% CI: 1.04, 2.34; p=0.033) higher odds of persistent SAM at discharge, respectively (**Figure 6F**; **Table S11**).

## DISCUSSION

Complicated SAM has typically been considered an immunosuppressed state, with functional responses to bacteria thought to reflect the immunoparalysis phenotype described in sepsis and multiple organ dysfunction syndrome(*31, 39, 40*). Previous pilot studies have found conflicting evidence for the capacity of innate immune cells to bind/phagocytose, upregulate activation markers, secrete immune mediators, initiate oxidative burst capacity, and kill bacteria in undernourished children(*33, 34*). By characterizing multiple domains of innate immune cell function simultaneously in an adequately-powered cohort, our data demonstrate that children who survive to hospital discharge achieve a compromise rather than a global deficiency in anti-bacterial innate immune cell function. We show that innate immune cells from children with SAM share some of the hallmark features of immunoparalysis, namely lower expression of HLA-DR and co-stimulatory molecules (CD86 and CD40) by monocytes, and lower pro-inflammatory cytokine production upon *in vitro* PAMP challenge (PCs 2-4). This accords with a previous study among 37 children admitted to the University Teaching Hospital, Lusaka with complicated SAM which found that blood dendritic cells (median: 0.51% IQR: 0.34–0.72 of peripheral blood mononuclear cells) had impaired IL-12 responses to LPS at hospital admission and a small percentage of children (17%; n=6) failed to upregulate HLA-DR upon LPS-stimulation(*41*). However, in parallel with lower monocyte activation/maturation and pro-inflammatory mediator secretion, we show that monocyte and neutrophil bacterial binding capacity (PC1 and 5) was markedly higher in the SAM group compared to the healthy control group. Given the association between elevated systemic pro-inflammatory mediators and mortality from complicated SAM(*15, 25, 26*), it is plausible that favoring bacterial containment via enhanced binding/phagocytosis over cytokine secretion by innate immune cells would provide a survival advantage during acute hospital admission. A similar adaptive balance has been observed for monocytes derived from adults hospitalized with gram-negative sepsis (n=3), which had lower expression of monocyte activation, pro-inflammatory cytokine and chemokine genes coinciding with enhanced phagocytosis and growth inhibition of live *E. coli in vitro* before versus after recovery(*27*). This monocyte ‘re-programming’ by sepsis favored angiogenesis and wound healing functions of septic monocytes *in vitro* which normalized in cells collected after recovery(*27*).

In this study we used a whole blood-based *in vitro* assay of bacterial binding capacity, including plasma components alongside all circulating cell types; the enhanced bacterial binding capacity that we observe in SAM could therefore reflect cell-intrinsic differences in activation, trafficking and expression of phagocytic receptors and/or increased levels of circulating bacterial binding proteins and opsonins that accelerate bacterial uptake. The SAM group had higher percentages of intermediate monocytes, which mature from classical monocytes and have higher expression of the Fcγ III receptor (CD16), which enhances phagocytosis and degranulation in response to IgG-opsonized bacteria(*42*); indirect evidence that monocytes from SAM have a more activated/mature phagocytic phenotype than those from healthy controls. Furthermore, concentrations of both LBP and sCD14, which are not phagocytic receptors but can bind gram-negative bacteria, mediate phagocytosis in some settings, and confer responsiveness to bacteria by binding to the membranes of non-monocytic (CD14-) cells in complex with LPS(*43–45*), were higher in plasma from children with SAM versus healthy controls. Recent evidence from experimental animal models supports our data that pathogen binding by innate immune cells is enhanced in the context of undernutrition; cutaneous monocytes from mice fed polynutrient deficient diets more readily internalized *Leishmania donovani* parasites and more *L. donovani*+ monocytes trafficked to the spleen relative to nutrient-sufficient animals(*46*). The mechanism/s by which bacterial binding capacity of innate immune cells is elevated in children with SAM and what impact this has on their subsequent trafficking to other tissues remains to be determined.

Moving beyond the view of SAM solely as an immunosuppressed/anergic state could support stratification of patients according to specific immunological risk factors; for example, a recent phase II double-blind trial of immune-based interventions among adults hospitalized with sepsis highlighted that clinical stratification based on hyper-inflammatory versus immunoparalysis phenotypes showed promise for identifying patients at highest mortality risk and guiding personalized immunotherapy (i.e. immune ‘boosting’ with recombinant IFNγ for patients with immunoparalysis or immune ‘dampening’ with Anakinra (a synthetic form of IL-1α receptor) for patients with macrophage activation-like syndrome)(*47*). Of 15 patients randomized to immunotherapy, 42.9% survived to 7 days with a decrease in sequential organ failure assessment score, relative to 10.0% of the 21 patients in the placebo arm(*47*). Notably, only 1 (0.01%) of our cohort of children with complicated SAM was septic at the time of blood sampling. Thus, whilst sepsis provides a pertinent analogue of the systemic endotoxin exposure observed in SAM and a ‘sepsis-like’ biomarker profile has been described among children recovering from SAM who die after hospital discharge(*25, 26*), SAM presents a distinct clinical and immunological condition.

It is unclear whether previously described differences in immune phenotype in children admitted with complicated SAM reflect nutritional status and/or specific patient characteristics/comorbidities. In the HOPE-SAM cohort, mortality over 1 year of post-discharge follow-up was associated with HIV infection (Adj. Hazard ratio (HR): 3.83; 95% CI: 2.15, 6.82), persistent SAM at discharge (Adj. HR: 2.28; 95% CI: 1.22, 4.25), cerebral palsy (Adj. HR: 5.60; 95% CI: 2.72, 11.50) and non-edematous SAM at admission (Adj. HR: 2.23; 95% CI: 1.24, 4.01)(*9*). Young age, SAM at discharge, non-edematous SAM at admission and cerebral palsy were also associated with hospital readmission or poor nutritional recovery after discharge(*6*). Symptoms of acute infection have previously been implicated in inpatient mortality among children hospitalized with SAM(*3*). We did not find evidence that these risk factors for poor clinical outcomes explained variation in scores for PC1 or PC5, the domains of immune function that best differentiated SAM cases from healthy controls. Thus, enhanced bacterial binding by innate immune cells from children with SAM relative to healthy controls is not solely due to their complications/comorbidities and may reflect as-yet-uncharacterized immune activating/regulatory processes specific to SAM. However, our data also demonstrate the relevance of demographic and clinical heterogeneity in children with SAM for other aspects of anti-bacterial innate immune cell function. PC2 was positively associated with MUAC, which reflects findings from a recent study among children admitted to hospitals in Kenya and Uganda with all-cause critical illness (46.0% were severely wasted). In that study, LPS-induced whole blood pro-inflammatory cytokine responses, including IL-6, IL-8 and TNFα, were lower among hospitalized versus healthy children in unadjusted analyses; this relationship was reversed after adjustment for MUAC(*38*), indicative of a distinct pattern of LPS-induced responses among wasted children. HIV profoundly impacts monocyte composition, trafficking and function, including maturation of monocytes into intermediate and non-classical sub-sets(*48*), circulating numbers due to increased homing to the gut(*49*), and release of sCD14 into circulation(*50*). HIV is also associated with the non-edematous form of SAM, a more severe pattern of wasting, increased risk of opportunistic infections, and HIV-driven inflammation is a risk factor for hospitalization with SAM among children living with HIV in LMIC(*36, 51, 52*). Consistent with HIV having an impact on the interrelationship between nutritional status and innate immune cell function, we found that excluding children with HIV strengthened the effect of SAM status on PC1 and PC5 in sensitivity analyses. HIV-SAM was also associated with lower monocyte activation in response to bacterial PAMP (PC3) than SAM alone. PC4 scores were lowest in the youngest age group suggesting that these children were less able to release MPO, an enzyme which catalyzes oxidative burst that is most abundantly expressed in neutrophil granules, upon bacterial PAMP stimulation. All children with SAM had higher basal MPO levels in unstimulated culture supernatants compared to healthy controls but basal levels were highest among children with SAM in the youngest age group (**Figure S7**), indicating that they had higher systemic oxidative stress and neutrophil degranulation relative to children in the older 2 age groups. Pre-exposure of monocytes from healthy adults to LPS *in vitro* (n=2) impairs their cytokine release, oxidative burst and pathogen killing compared to saline-treated monocytes (n=3)(*53*). Thus, whilst higher basal levels of MPO may promote pathogen killing by young children with SAM in the short-term, subsequent deficits in pre-formed MPO granules and capacity for MPO release by granulocytes may compromise their defense against infections in the longer term(*53*). We did not find evidence that coincident symptoms of infection influenced any of the immune function PCs. This was an unexpected finding given that acute infections expand and prime immune cells *in vivo* and 60.0% of our cohort had at least 1 symptom of infection at the time of immune function assessment. However, since we were reliant on physician examination-based identification of infections per standard of care at the hospital recruitment sites, our data likely underestimated the prevalence of infections. It is also plausible that reduced cytokine production and monocyte activation marker expression in response to bacterial PAMP might mask infectious symptoms in children with SAM. Inclusion of molecular diagnostics in future studies would provide greater scope to characterize the impact of concurrent infections and specific pathogens on SAM immune function.

Nutritional rehabilitation after hospital admission relies on resolution of clinical complications alongside therapeutic re-feeding; an outstanding question which we are pursuing using longitudinal samples from this cohort during post-discharge follow-up, is whether or not rehabilitation of immune cell function accords with anthropometric recovery(*36*). Cross-sectional evaluation during inpatient rehabilitation demonstrated that bacterial binding (PC1) and neutrophil responses (PC5) were negatively associated with the duration between hospital admission and immune function assessment. High scores for PC1 and PC5 differentiated children with SAM from healthy controls, therefore their negative relationship with inpatient treatment duration in the SAM group is indicative of these responses ‘normalizing’ towards those seen in healthy immune cells. This has previously been suggested in pilot studies of blood immune cell function (n=10)(*54*), and for circulating dendritic cells from children hospitalized with SAM in Zambia which were higher in number, LPS-induced HLA-DR expression, and unstimulated IL-12 secretion at discharge than at hospital admission (n=27)(*41*). Normalization of innate immune cell function towards that seen in healthy children could reflect and contribute to resolution of other physiological features disrupted in complicated SAM and posited as drivers of systemic inflammation (e.g. dysbiosis, metabolic profiles, gut barrier function)(*26, 55–57*). For example, pro-inflammatory biomarkers that discriminated between the plasma proteome of 30 children with SAM and 21 healthy children in Bangladesh normalized following 4 weeks of a microbiota-directed legume-based feeding intervention in concert with increased WHZ and microbiota maturity(*55*). The ‘immune rehabilitation’ of anti-bacterial responses was incomplete in our cohort of SAM inpatients since all but 9/126 (7.1%) had PC1 scores greater than the median score for the healthy group. Functional indicators of monocyte activation and mediator secretion in response to PAMP (PC2-4) were unaffected by time to immune assay, consistent with their stronger association with more stable participant characteristics (age group, HIV status) and continuous relationship with MUAC rather than with SAM status.

The clinical importance of normalizing anti-bacterial innate immune cell function as part of rehabilitation from SAM is supported by evidence that PC1 scores during hospitalization were associated with persistent SAM status at discharge, a predictor of both mortality and persistent wasting over a year of post-discharge follow-up in HOPE-SAM(*6, 9*). Scores for PC3 (monocyte activation), which tended to be lower in children with SAM than healthy controls, were associated with higher odds of persistent wasting within the SAM group. Given the energetic cost of monocyte activation and turnover(*58*), which is associated with changes in circulating lipid levels(*58*), it is plausible that heightened pro-inflammatory capacity of blood monocytes might benefit anti-bacterial defense but compromise weight gain in children recovering from SAM. The association between immune cell function and wasting that we identified accords with evidence from HIV-negative children recovering from SAM in Kenya for whom MUAC and WHZ gains over the first 60 days post-discharge were negatively associated with circulating biomarkers of systemic inflammation(*56*).

Whilst this study advances our understanding of immune function in SAM, our data should be interpreted in the context of several limitations. Firstly, our cohort included only children able to provide a ≥2mL blood sample, necessarily excluding the most acutely unwell children and those who died prior to enrolment or blood sampling; our findings are therefore most applicable to children who survive to hospital discharge. Second, we undertook immune function assessment across a range of time-points after hospital admission which contributed to immune heterogeneity in the SAM group. Adjusting analyses for time to immunoassay, presenting cross-sectional analyses of PCs by time to immunoassay and sensitivity analyses focusing on children for whom immune function was assessed prior to the day of discharge, go some way to demonstrate the effect of this variable on our findings and the generalizability of the SAM-related differences that we see despite this limitation. Thirdly, there remains much to learn about the immunobiology of SAM(*24, 34*), much of which was not possible to explore in this study but would be pertinent to our interpretation. We chose to focus on gram-negative bacterial PAMP which are among the most common clinical isolates from children with SAM but unlikely to be the only group of infections nor LPS the only PAMP translocated in the context of SAM; in our cohort alone, 11.5% of the SAM group had fungal infections (oral candidiasis) and 13.8% tuberculosis at the time of their immunoassay, and gram-positive bacteria, viruses, fungi and parasites are also prevalent in undernourished children(*10, 15*). Similarly, SAM is implicated in impaired adaptive immunity (*33, 59*), and, although we provide some evidence for bulk differences in bacterial binding by Lin+ lymphocyte populations in children with SAM, detailed analyses of adaptive immunity was beyond our scope. Fourth, though our study is the largest of its kind and adequately-powered for our primary comparison between the SAM and healthy groups, it is increasingly apparent from clinical studies of SAM, including our own(*6, 9, 51*), that specific co-morbidities contribute to sub-conditions within current SAM diagnostic criteria (e.g. HIV-SAM, cerebral palsy-SAM, edematous/non-edematous SAM); a larger or comorbidity-focused sample size would be required to conclusively evaluate the impact of these variables on immune function. Since some of these comorbidities are shared by hospitalized children without SAM and recent studies suggest that these children have distinct anti-bacterial pro-inflammatory mediator responses relative to healthy controls to those we observe in SAM(*38*), it would be desirable to include such children as an additional comparison group in future studies.

Overall, we provide evidence for a functional shift in how innate immune cells respond to bacterial PAMP during hospitalization with complicated SAM, with enhanced bacterial binding capacity of monocytes and neutrophils differentiating children with SAM from healthy controls alongside characteristics of impaired monocyte maturation and pro-inflammatory cytokine secretion to bacterial PAMP. Differences in bacterial binding capacity are not explained by patient demography or clinical comorbidities, but are more tractable to inpatient treatment than other domains of innate immune function and contribute to persistent SAM at discharge; a predictor of mortality and poor nutritional recovery after hospital discharge(*6, 9*). Thus, innate immune cells from children with SAM strike a balance in their response to new infectious challenges, which impacts inpatient rehabilitation and may contribute to persistent health deficits after hospital discharge.

## MATERIALS AND METHODS

### Study design

<u>IMMUN</u>e function <u>O</u>f children with <u>SAM</u> (IMMUNO-SAM) is a translational immunology study designed to characterize anti-bacterial innate immune cell function using blood samples from children under 5 years old hospitalized with complicated SAM at three tertiary referral hospitals in Lusaka (University Teaching Hospital), Zambia and Harare (Harare Central Hospital and Parirenyatwa Hospital), Zimbabwe. All participants were enrolled into <u>H</u>ealth <u>O</u>utcomes and <u>P</u>athog<u>E</u>nesis of <u>SAM</u> (HOPE-SAM), a longitudinal observational study for which the primary outcome was mortality over 1 year of post-discharge follow-up; clinical study protocols and outcomes are published elsewhere(*6, 9, 36, 60*).

Ethical approval was obtained from the University of Zambia Biomedical Research Ethics Committee, and the Medical Research Council of Zimbabwe. The ethics committee of the Queen Mary University of London provided an advisory review. Caregivers of all participants provided written informed consent for their participation.

### Study participants

***SAM cases*** were children aged 0-59 months, admitted to one of the three hospital sites with complicated SAM. SAM was diagnosed according to the current WHO definition as WHZ <−3, MUAC < 115 mm and/or the presence of nutritional edema for children above 6 months of age, or WHZ <−3 and/or nutritional edema for those aged under 6 months(*1*). ***Healthy controls*** were children aged 6-59 months who were adequately-nourished (WHZ >– 1) and clinically well (no symptoms of acute illness or current infections). Additional inclusion criteria were: 1) enrolled into HOPE-SAM after ethical approval of IMMUNO-SAM procedures (August 2017); 2) known HIV status; 3) ≥2mL blood sample collected during hospitalization (SAM group) or at baseline visit (healthy controls); and, 4) no underlying chronic gastrointestinal disease or a known malignancy. For SAM cases, any blood sample collected between their initial clinical assessment at baseline and their discharge from hospital was admissible to the study. Age groups were pre-specified for the HOPE-SAM study (0-5 mo, 6-11 mo, 12-23 mo and 24-59 mo)(*36*); the 7 children in the SAM group who were aged under 6 mo at admission were included in the youngest age group (0-11 mo).

### Clinical procedures

Clinical, household and demographic characteristics from the primary caregiver of each child via questionnaire at study baseline, i.e. as soon as possible after hospital admission. Clinical care for SAM cases was implemented according to country-specific guidelines based on WHO guidelines(*1, 2*). Where HIV status could not be confirmed by the clinical team, HIV status for mothers and for children aged >18mo was determined by the HOPE-SAM study team using a rapid antibody test algorithm, and for children <18mo using HIV DNA PCR. Daily clinical review of each child with SAM was undertaken by a study physician using a standardized inpatient review form. Anthropometry was conducted at baseline and repeated for SAM cases on the day of discharge. Weight was recorded to the nearest 10g using a Seca384 electronic scale for infants and to the nearest 100g using a Seca874 electronic scale for older children. MUAC was recorded to the nearest 1mm using WHO/UNICEF MUAC tape. Length/height were measured to the nearest 0.1cm using UNICEF measuring boards.

Children were discharged to outpatient community-based management of nutritional rehabilitation according to WHO-adapted country guidelines, which specify that discharge is appropriate when a child’s medical complications, including edema, are resolving, they are clinically well, alert and their appetite has returned(*1, 2*).

### Blood sample collection

Blood samples (1 mL/kg up to a maximum of 5.4 mL) were collected into endotoxin-free EDTA-treated blood collection tubes. Blood was not collected from children with severe anemia (known hemoglobin <60 g/L).

### Bacterial binding assay

On the day of blood sample collection, 100ul of undiluted whole blood was added to pre-prepared cryovials with: culture media only (RPMI 1640 medium supplemented with 1% v/v penicillin-streptomycin; unstimulated control), and 5×10^5^ AF488-conjugated *E. coli* bioparticles diluted in culture medium. Cultures were mixed thoroughly and incubated for 1h at 37⁰C, 6% CO2 using the CO2Gen compact system. After 1h, cultured cells were treated with 1× FACS lysing and fixative buffer for 10mins, washed in phosphate-buffered saline (PBS), resuspended in cell freezing medium (PBS with 10% v/v dimethyl sulfoxide (DMSO), 40% v/v heat-inactivated fetal bovine serum (FBS)), and gradually frozen overnight at 1°C/min to -80⁰C using a Mr. Frosty freezing container. Details of key reagents are provided in **Table S12.**

### Whole blood cultures

On the day of blood sample collection, 750ul of whole blood was diluted in 1.45mL culture media. 500ul/well of diluted blood was added to wells of pre-prepared 48-well culture plates containing culture media alone (unstimulated control), HKST (final concentration: 1×10^8^cells/mL) and ultrapure *E. coli* LPS (final concentration: 0.25EU/mL). Whole blood cultures were incubated for 24h at 37⁰C, 6% CO2 using the CO2Gen compact system. Cell-free culture supernatants were harvested and cryopreserved at -80⁰C. Cultured blood cells were subjected to red blood cell lysis, fixed and cryopreserved, as above. Details of key reagents are provided in **Table S12**

### Plasma, stool and culture supernatant ELISA

Biomarkers of systemic inflammation (C-reactive protein (CRP), monocyte/macrophage activation (soluble CD14 (sCD14), sCD163) and LPS-binding protein (LBP) were quantified in plasma (Biotechne Quantikine ELISA kits) and myeloperoxidase (MPO; Immundianostik, Bensheim, Germany) and neopterin (GenWay Biotech Inc, San Diego, CA, USA) in stool via ELISA at Zvitambo and TROPGAN laboratories; methods previously reported(*6, 9, 36*).

TNFα, IL-6, IL-8 and MPO concentrations were quantified in whole blood culture supernatants by ELISA (reagent details in **Table S12**) at Zvitambo and TROPGAN laboratories. ELISA lower limits of detection were: TNFα - 15.6pg/ml, IL-6 - 4pg/ml, IL-8 -31.2pg/ml and MPO - 62.5pg/ml. Concentrations below the assay detection limit were censored at the limit of detection; those greater than the top standard concentration were re-run at higher dilution and multiplied by the dilution factor.

### Flow cytometry

Cryopreserved bacterial binding assay cell samples were labelled with the following fluorochrome-labelled antibody panel: Lin (CD3, CD19, CD20, CD56)-APC, CD66b-PerCPCy5.5, CD16-APCCy7, HLA-DR-PECy7, CD14-PE; **Table S12**) to identify which cell types were bound to the AF488-labelled *E. coli*-coated bioparticles. Cryopreserved whole blood culture cell samples were labelled with the following fluorochrome-labelled antibody panel: Lin (CD3, CD19, CD20, CD56)-APC, CD66b-APC, CD16-APCCy7, HLA-DR-PECy7, CD14-PE, CD86-FITC, CD40-PerCPCy5.5; **Table S12**). Flow cytometry was conducted on the day of antibody labelling using 6-colour BD Biosciences FACSVerse cytometers (488nm and 633nm lasers) at Zvitambo and TROPGAN laboratories after daily instrument calibration using CS&T beads and the Performance QC program in FACSuite software. Flow cytometry gating strategies (**Figure S2**) were implemented in FlowJo version 10.8.1 (FlowJo, LLC, USA); universal gates were based on fluorescence-minus-one controls for each marker, applied to all samples, and adjusted per participant based on their corresponding assay-specific unstimulated control sample.

### Statistical Analyses

Proportional data were analyzed by fractional regression, mean (*E. coli*-AF488) and median (HLA-DR, CD86 and CD40) fluorescence intensity (FI) data were left-censored at 1 and analyzed by tobit regression after log-transformation, and, soluble mediator concentrations in culture supernatants were left-censored at the assay limit of detection and analyzed by tobit regression after log-transformation.

LPS- and HKST-induced median fluorescence intensities (HLA-DR, CD86 and CD40) and soluble mediator concentrations were calculated for each participant by subtracting values for their corresponding unstimulated cultures, left-censored at 0 (i.e. no detectable response above that in unstimulated cultures). There was no background meanFI or proportions of *E. coli*+ cells in unstimulated cultures, these were uncensored. The effect of SAM status on log-transformed proportions of *E. coli*+ cells was analyzed by fractional regression, on log-transformed LPS- and HKST-induced monocyte medianFI (HLA-DR, CD86 and CD40) and log-transformed LPS- and HKST-induced supernatant mediator concentrations by tobit regression, and on log-transformed *E. coli*-AF488 meanFI by linear regression. tSNE plots for all compensated cell surface markers (Lin, CD14, CD16, CD66b, HLA-DR) were generated for 500 *E. coli*+ leukocytes events per participant for all SAM cases and healthy controls and FlowSOM analysis applied using the cytofkit2 package in R.

Log-transformed PAMP-induced innate immune cell function outcome measures for all SAM cases (n=126) and healthy controls (n=73) with complete data were standardized (mean=0, standard deviation=1) and entered into a single principal components analysis. Resulting principal components (PCs) with an eigenvalue >1 and ≥1 of the original variables loading onto the PC with a factor loading ≥±0.3 were selected for subsequent analyses. The effect of SAM status on immune profiles (PC1-5 scores) was analyzed by linear regression before and after adjusting for age group, sex, HIV status, hospital site and immune function assessment on the day of discharge. Variables were selected for inclusion in adjusted model based on causal inference methodology using a directed acyclic graph (DAG) of exposure and outcome measures generated using Daggity software(*61*) (**Figure S8A**). We undertook pre-specified sensitivity analyses excluding: 1) HIV-positive participants and 2) participants for whom immune function was assessed on the same day as discharge.

The effects of clinically-relevant demographic and clinical characteristics on anti-bacterial innate immune function were estimated for each PC using cross-fit partialling-out lasso linear regression with default lambda selection. Sex, hospital site, time to immunoassay, baseline WHZ, baseline HAZ, birthweight and cerebral palsy status were offered as control variables based on their putative contribution to variation in both immune function and clinical outcomes (**Figure S8B**).

To characterize the cross-sectional relationship between immune function and duration of hospitalization, we analyzed PC1-5 scores by time to immunoassay via linear regression, adjusting for hospital site, symptoms of infection on the day of immune function assessment and immune function assessment on the day of discharge (**Figure S8C**).

We tested the effect of immune function assessed prior to the day of discharge on SAM status at discharge via logistic regression, adjusting for sex, baseline age group, HIV status, hospital site, time to immunoassay, and baseline MUAC (**Figure S8D**). Children discharged against medical advice were excluded from these analyses.

All statistical analyses were performed using STATA version 17 (StataCorp LLC, USA). Outcome variables were plotted using Prism version 9 (GraphPad Software Inc., USA) and R version 4.2.0.

## Data Availability

Requests for further information, resources and reagents should be directed to the study Principle Investigator, Claire D. Bourke. Participant samples were collected in Zambia and Zimbabwe under local ethical approval and consenting procedures and are therefore restricted my Material Transfer Agreements.

## ACKNOWLEDGMENTS

We thank all the caregivers and children who participated in the HOPE-SAM study and the staff at Harare Central Hospital, Parirenyatwa Hospital and the University Teaching Hospital for hosting the study team. We acknowledge the wider HOPE-SAM study team (members listed here: https://bmjopen.bmj.com/content/9/1/e023077) and Stephen Moyo (logistics), Philippa Rambanepasi (finance), Virginia Sauramba and Miyoba Chipunza (compliance), and Agatha Muyenga (administration), in particular, for their additional support for this study at Zvitambo and TROPGAN. We acknowledge Emma Chambers for initial support with tSNE.

## AUTHOR CONTRIBUTIONS

Implementation of laboratory work: TNP, SR, MG, PM, SM, TH, TR, LC, CDB Laboratory management: KM, KZ

Data processing: CDD, JT, CDB, JPS, RCR, MB Data management: RN

Clinical data collection: MB, FDM, DN, KC, CK, BM

HOPE-SAM design and clinical management: BA, MB, PK, AJP, KN Statistical review: MS, RN

Study design: CDB Statistical analysis: CDB Drafted manuscript: CDB

Reviewed manuscript and approved submission: all co-authors

## REFERENCES

1. WHO, in Guideline: Updates on the management of severe acute malnutrition in infants and children. (World Health Organization Copyright © World Health Organization 2013., Geneva, 2013).

2. G. WHO, Management of the child with a serious infection or severe malnutrition: guidelines for care at the first-referral level in developing countries. (2000).

3. R. Karunaratne, J. P. Sturgeon, R. Patel, A. J. Prendergast, Predictors of inpatient mortality among children hospitalized for severe acute malnutrition: a systematic review and meta-analysis. The American journal of clinical nutrition, (2020).

4. UNICEF, WHO, W. Bank, “Levels and trends in child malnutrition: UNICEF/WHO/The World Bank Group joint child malnutrition estimates: key findings of the 2021 edition,” (2021).

5. D. Sharrow et al., Global, regional, and national trends in under-5 mortality between 1990 and 2019 with scenario-based projections until 2030: a systematic analysis by the UN Inter-agency Group for Child Mortality Estimation. The Lancet Global Health 10, e195–e206 (2022).

6. M. Bwakura-Dangarembizi et al., Recovery of children following hospitalisation for complicated severe acute malnutrition. Maternal & Child Nutrition 18, e13302 (2022).

7. M. Kerac et al., Follow-up of post-discharge growth and mortality after treatment for severe acute malnutrition (FuSAM study): a prospective cohort study. PLOS ONE 9, e96030 (2014).

8. A. Talbert et al., Diarrhoea complicating severe acute malnutrition in Kenyan children: a prospective descriptive study of risk factors and outcome. PLOS ONE 7, e38321 (2012).

9. M. Bwakura-Dangarembizi et al., Risk factors for postdischarge mortality following hospitalization for severe acute malnutrition in Zimbabwe and Zambia. The American journal of clinical nutrition 113, 665–674 (2021).

10. J. A. Berkley et al., Daily co-trimoxazole prophylaxis to prevent mortality in children with complicated severe acute malnutrition: a multicentre, double-blind, randomised placebo-controlled trial. The Lancet. Global health 4, e464–473 (2016).

11. M. Ngari et al., Mortality during and following hospital admission among school-aged children: a cohort study. Wellcome Open Research 5, 234 (2021).

12. M. M. Ngari et al., Changes in susceptibility to life-threatening infections after treatment for complicated severe malnutrition in Kenya. The American journal of clinical nutrition 107, 626–634 (2018).

13. J. A. Berkley et al., Bacteremia among children admitted to a rural hospital in Kenya. N Engl J Med 352, 39–47 (2005).

14. A. L. Page et al., Infections in children admitted with complicated severe acute malnutrition in Niger. PLoS One 8, e68699 (2013).

15. S. Attia et al., Mortality in children with complicated severe acute malnutrition is related to intestinal and systemic inflammation: an observational cohort study. The American journal of clinical nutrition 104, 1441–1449 (2016).

16. K. Maitland et al., Children with severe malnutrition: can those at highest risk of death be identified with the WHO protocol? PLOS Medicine 3, e500 (2006).

17. I. Olofin et al., Associations of suboptimal growth with all-cause and cause-specific mortality in children under five years: a pooled analysis of ten prospective studies. PLoS One 8, e64636 (2013).

18. B. Amadi et al., Adaptation of the small intestine to microbial enteropathogens in Zambian children with stunting. Nature Microbiology 6, 445–454 (2021).

19. S. Mero et al., Prevalence of diarrhoeal pathogens among children under five years of age with and without diarrhoea in Guinea-Bissau. PLoS Neglected Tropical Diseases 15, e0009709 (2021).

20. G. Patterson et al., Environmental, metabolic, and inflammatory factors converge in the pathogenesis of moderate acute malnutrition in children: an observational cohort study. The American Journal of Tropical Medicine and Hygiene, (2021).

21. J. Lauer et al., Markers of environmental enteric dysfunction are associated with poor growth and iron status in rural ugandan infants. The Journal of nutrition 150, (2020).

22. S. Syed et al., Serum anti-flagellin and anti-lipopolysaccharide immunoglobulins as predictors of linear growth faltering in Pakistani infants at risk for environmental enteric dysfunction. PLoS ONE 13, e0193768 (2018).

23. K. D. J. Jones et al., Mesalazine in the initial management of severely acutely malnourished children with environmental enteric dysfunction: a pilot randomized controlled trial. BMC Medicine 12, 133 (2014).

24. C. D. Bourke, J. A. Berkley, A. J. Prendergast, Immune Dysfunction as a Cause and Consequence of Malnutrition. Trends in Immunology 37, 386–398 (2016).

25. J. M. Njunge et al., Biomarkers of post-discharge mortality among children with complicated severe acute malnutrition. Scientific Reports 9, 5981 (2019).

26. B. Wen et al., Systemic inflammation and metabolic disturbances underlie inpatient mortality among ill children with severe malnutrition. Science Advances 8, eabj6779 (2022).

27. I. N. Shalova et al., Human monocytes undergo functional re-programming during sepsis mediated by hypoxia-inducible factor-1alpha. Immunity 42, 484–498 (2015).

28. P. M. de Azambuja Rodrigues et al., Proteomics reveals disturbances in the immune response and energy metabolism of monocytes from patients with septic shock. Sci Rep 11, 15149 (2021).

29. T. Akoumianaki et al., Uncoupling of IL-6 signaling and LC3-associated phagocytosis drives immunoparalysis during sepsis. Cell Host & Microbe 29, 1277–1293.e1276 (2021).

30. S.-C. Cheng et al., Broad defects in the energy metabolism of leukocytes underlie immunoparalysis in sepsis. Nat Immunol 17, 406–413 (2016).

31. J. Leentjens et al., Reversal of immunoparalysis in humans in vivo. American Journal of Respiratory and Critical Care Medicine 186, 838–845 (2012).

32. G. Monneret et al., Marked elevation of human circulating CD4 + CD25+ regulatory T cells in sepsis-induced immunoparalysis. Crit Care Med. 31, (2003).

33. M. J. Rytter, L. Kolte, A. Briend, H. Friis, V. B. Christensen, The immune system in children with malnutrition - a systematic review. PLoS ONE 9, e105017 (2014).

34. C. D. Bourke, K. D. J. Jones, A. J. Prendergast, Current understanding of innate immune cell dysfunction in childhood undernutrition. Frontiers in Immunology 10, (2019).

35. A.-L. Page et al., Infections in children admitted with complicated severe acute malnutrition in Niger. PLOS ONE 8, e68699 (2013).

36. M. Bwakura-Dangarembizi et al., Health outcomes, pathogenesis and epidemiology of severe acute malnutrition (HOPE-SAM): rationale and methods of a longitudinal observational study. BMJ Open 9, e023077 (2019).

37. A. A. Patel et al., The fate and lifespan of human monocyte subsets in steady state and systemic inflammation. The Journal of Experimental Medicine 214, 1913–1923 (2017).

38. L. S. Uebelhoer et al., Toll-like receptor-induced immune responses during early childhood and their associations with clinical outcomes following acute illness among infants in sub-Saharan Africa. Frontiers in Immunology 12, (2022).

39. T. F. Manzoli et al., Lymphocyte count as a sign of immunoparalysis and its correlation with nutritional status in pediatric intensive care patients with sepsis: A pilot study. *Clinics (Sao Paulo*, Brazil) 71, 644–649 (2016).

40. M. W. Hall et al., Immunoparalysis and nosocomial infection in children with multiple organ dysfunction syndrome. Intensive Care Med 37, 525–532 (2011).

41. S. M. Hughes et al., Dendritic cell anergy results from endotoxemia in severe malnutrition. The Journal of Immunology 183, 2818–2826 (2009).

42. S. Gordon, Phagocytosis: an immunobiologic process. Immunity 44, 463–475 (2016).

43. R. S. Jack, U. Grunwald, F. Stelter, G. Workalemahu, C. Schütt, Both membrane-bound and soluble forms of CD14 bind to gram-negative bacteria. European journal of immunology 25, 1436–1441 (1995).

44. U. Grunwald et al., Monocytes can phagocytose Gram-negative bacteria by a CD14-dependent mechanism. J Immunol 157, 4119–4125 (1996).

45. E. A. Frey et al., Soluble CD14 participates in the response of cells to lipopolysaccharide. J Exp Med 176, 1665–1671 (1992).

46. E. Osorio et al., Malnutrition-related parasite dissemination from the skin in visceral leishmaniasis is driven by PGE2-mediated amplification of CCR7-related trafficking of infected inflammatory monocytes. PLoS Neglected Tropical Diseases 17, e0011040 (2023).

47. K. Leventogiannis, et al., Toward personalized immunotherapy in sepsis: The PROVIDE randomized clinical trial. Cell Reports Medicine 3, (2022).

48. P. Chen et al., Perturbations of monocyte subsets and their association with T helper cell differentiation in acute and chronic HIV-1-infected patients. Frontiers in Immunology 8, 272 (2017).

49. K. Allers et al., Macrophages accumulate in the gut mucosa of untreated HIV-infected patients. The Journal of Infectious Diseases 209, 739–748 (2013).

50. A. J. Prendergast et al., Intestinal damage and inflammatory biomarkers in Human Immunodeficiency Virus (HIV)–exposed and HIV-infected Zimbabwean infants. The Journal of Infectious Diseases 216, 651–661 (2017).

51. A. Prendergast et al., Hospitalization for severe malnutrition among HIV-infected children starting antiretroviral therapy. *AIDS (London*, England) 25, 951–956 (2011).

52. A. J. Prendergast et al., Inflammatory biomarkers in HIV-infected children hospitalized for severe malnutrition in Uganda and Zimbabwe. *AIDS (London*, England) 33, 1485–1490 (2019).

53. I. Grondman et al., Endotoxin-induced immunotolerance is associated with loss of monocyte metabolic plasticity and reduction of oxidative burst. Journal of Leukocyte Biology 106, (2019).

54. R. Peixoto Paes-Silva, M. Correia de Macedo É, M. T. Oliveira Tomiya, C. M. Machado Barbosa de Castro, Immune respose of severe malnutrition children treated according to the protocol of the World Health Organisation. Nutricion hospitalaria 32, 638–644 (2015).

55. J. L. Gehrig et al., Effects of microbiota-directed foods in gnotobiotic animals and undernourished children. Science 365, (2019).

56. J. Njunge et al., Systemic inflammation is negatively associated with early post discharge growth following acute illness among severely malnourished children - a pilot study. Wellcome Open Research 5, 248 (2021).

57. N. Calder et al., Modifying gut integrity and microbiome in children with severe acute malnutrition using legume-based feeds (MIMBLE): A pilot trial. *Cell reports*. Medicine 2, 100280 (2021).

58. V. K. Patel, H. Williams, S. C. H. Li, J. P. Fletcher, H. J. Medbury, Monocyte inflammatory profile is specific for individuals and associated with altered blood lipid levels. Atherosclerosis 263, 15–23 (2017).

59. N. Nabukeera et al., Thymus size and its correlates among children admitted with severe acute malnutrition: a cross-sectional study in Uganda. BMC Pediatrics 21, (2021).

60. M. Bwakura-Dangarembizi et al., Fat and lean mass predict time to hospital readmission or mortality in children treated for complicated severe acute malnutrition in Zimbabwe and Zambia. The British journal of nutrition, 1–25 (2022).

61. J. Textor, B. van der Zander, M. K. Gilthorpe, M. Liskiewicz, G. T. H. Ellison, Robust causal inference using directed acyclic graphs: the R package ‘dagitty’. International Journal of Epidemiology 45, 1887–1894 (2016).

